# Epidural stimulation of the cervical spinal cord improves voluntary motor control in post-stroke upper limb paresis

**DOI:** 10.1101/2022.04.11.22273635

**Authors:** Marc P. Powell, Nikhil Verma, Erynn Sorensen, Erick Carranza, Amy Boos, Daryl Fields, Souvik Roy, Scott Ensel, Beatrice Barra, Jeffery Balzer, Jeff Goldsmith, Robert M. Friedlander, George Wittenberg, Lee E. Fisher, John W Krakauer, Peter C. Gerszten, Elvira Pirondini, Douglas J Weber, Marco Capogrosso

**Author notes:** Co-first authors (Marc P. Powell, Nikhil Verma, Erynn Sorensen). Co-senior authors (Elvira Pirondini, Douglas J Weber, Marco Capogrosso).

## Abstract

A large proportion of cerebral strokes disrupt descending commands from motor cortical areas to the spinal cord which can results in permanent motor deficits of the arm and hand^1,2^. However, below the lesion, the spinal circuits that control movement^5^ remain intact and could be targeted by neurotechnologies to restore movement^6–9^. Here we demonstrate that by engaging spinal circuits with targeted electrical stimulation we immediately improved voluntary motor control in two participants with chronic post-stroke hemiparesis. We implanted a pair of 8-contact percutaneous epidural leads on the lateral aspect of the cervical spinal cord to selectively target the dorsal roots that provide excitatory inputs to motoneurons controlling the arm and hand^10,11^. With this strategy, we obtained independent activation of shoulder, elbow and hand muscles. Continuous stimulation through selected contacts at specific frequencies enabled participants to perform movements that they had been unable to perform for many years. Overall, stimulation improved strength, kinematics, and functional performance. Unexpectedly, both participants retained some of these improvements even without stimulation, suggesting that spinal cord stimulation could be a restorative as well as an assistive approach for upper limb recovery after stroke.

## INTRODUCTION

Globally, 1 in 4 people will suffer from a stroke^12^. Of these people, nearly three quarters will exhibit lasting deficits in motor control of their arm and hand^4^ leading to enormous personal and societal impact^13^. One reason for this is that physical therapy in the acute phase, the current gold-standard, unfortunately does not significantly reduce upper limb impairment^14^.

Patients with chronic stroke exhibit a stereotypical motor syndrome of the upper limb that can be decomposed in independently quantifiable deficits^1^: loss of strength, reduced dexterity, intrusion of aberrant synergies and spasticity. This *paresis* phenotype emerges from damage to the cortico-spinal tract (CST), which disrupts connections between the cortex and the cervical spinal circuits controlling arm and hand movements^1,3,15^. In most cases this damage is incomplete, but the spared CST is not sufficient to generate functional movement.

We conjectured that voluntary motor control could be restored by amplifying the latent capacities of the damaged CST. Specifically, we hypothesized that modulating the excitability of intact sub-lesional spinal circuits would increase their responsiveness to remaining CST neurons thereby restoring the ability of these supra-spinal inputs to drive movement. One pathway for the manipulation of spinal circuit excitability is through stimulation of primary sensory afferent neurons within the dorsal roots of the spinal cord. Indeed, a century of research has shown that primary afferent neurons are a major source of excitatory drive for motoneurons and other spinal circuits^7– 9,16–20^. We and others showed that epidural spinal cord stimulation (SCS), a clinically approved technology, can be used to directly recruit these afferents^10,21,22^ thus unlocking a realistic path to test our hypothesis in humans with chronic stroke.

Clinical support for this approach comes from studies exploring the use of SCS for the recovery of locomotion after spinal cord injury^23–29^ (SCI). While most of these studies focused on quantifying the cyclic patterns of movement involved in walking^25,27,30^, several groups reported that SCS enabled people with complete leg paralysis to produce voluntary single joint movements^23,24,26^. This facilitation of motor intent was immediate and required SCS to remain on; an assistive effect^23,24,28^. Importantly, this assistive effect is conceptually different from technologies like functional electrical stimulation (FES) which produce involuntary movements^31,32^. SCS does not produce movement, instead it facilitates the ability of residual proprio-spinal and supraspinal inputs to activate spinal motoneurons, thereby enabling volitional movement^6,24,33,34^. Moreover, when the assistive effect of SCS is combined with prolonged training involving physical activity, it promoted long-lasting recovery of voluntary leg motor function even in the absence of stimulation; a therapeutic effect^26,29^.

Despite these encouraging findings motor recovery in the leg, translation of these results to post-stroke upper-limb motor syndrome is hindered by significant scientific and technical challenges. Dexterous control of the arm and hand relies more heavily on cortico-spinal input^35^ than does control of locomotion^19,36^, so for the same amount of remaining CST, the degree of spinal circuit potentiation required may be greater. Moreover, the physiology of post-stroke motor deficits is different than SCI and therefore the response to SCS may be qualitatively different. For example, spasticity in stroke responds differently to pharmacological treatments that are effective in SCI^37^. Finally, from a technical perspective, the cervical enlargement is significantly longer and larger than the lumbosacral spinal cord which makes current clinical paddle leads insufficient to cover all the segments innervating upper limb muscles^10^. We addressed this with a neurosurgical approach that implants two staggered linear leads on the lateral aspect of the cervical cord to target each dorsal root innervating arm and hand muscles at their entry into the spinal cord^8,10,38^. We devised a battery of scientific and clinical assessments to verify if SCS could improve cortico-spinal control and quantify the assistive effects of SCS on strength, dexterity, synergies, and spasticity.

## RESULTS

### Experimental framework to quantify the effects of SCS on post-stroke motor hemiparesis

We recruited two participants with chronic post-stroke motor deficits to evaluate the effects of cervical SCS on motor performance (**Figure 1**). SCS01 suffered a ischemic stroke while SCS02 suffered a hemorrhagic stroke >5 years and >1 year prior to study enrollment, respectively. SCS01’s lesion was localized to the internal capsule, midbrain, and pons, while SCS02’s lesion was larger, affecting a large percentage of the corona radiata of the right hemisphere (**Figure 1d, Extended Data Figure 1)**. In both cases, extensive damage to the CST resulted in chronic upper limb impairment. We performed a tractography analysis using high-definition fiber tracking (HDFT) to compare the integrity of CST axons between the lesioned and healthy hemispheres of both participants (**Extended Data Figure 1**). We measured relative white matter integrity by comparing Fractional anisotropy (FA) of the lesion hemisphere against the non-lesioned hemisphere and obtained FAS=0.17 for SCS01 and FAS=0.35 for SCS02 and (FAS=0 no impairment, see methods) indicating severe unilateral damage to the CST. This was reflected in pre-study Fugl-Meyer motor assessments of 35/66 (SCS01) and 15/66 (SCS02), indicative of moderate and severe impairment respectively.

**Figure 1.**
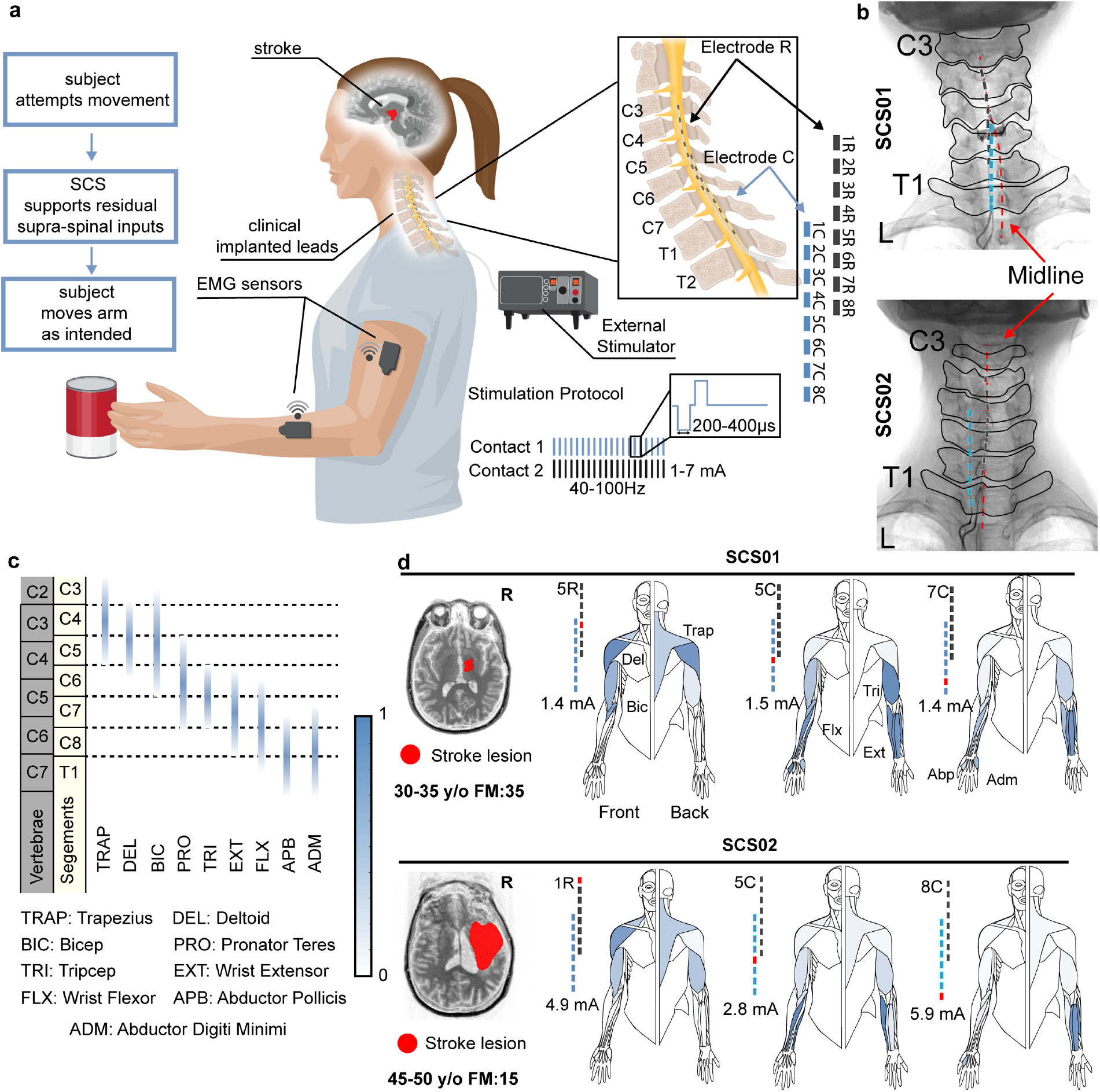
Experimental framework and stimulation specificity. **(a)** Schematic of the experimental platform and paradigm. While participants performed an upper limb motor task, we measured wireless electromyographic (EMG) activity from muscles of the arm and hand. We delivered electrical stimulation to the cervical spinal cord via two 8-cont leads (Rostral, R; Caudal, C) implanted in the cervical spinal cord. Stimulation through selected contacts simultaneously was controlled via percutaneous connections using an external stimulator. **(b)** X-rays of both participants showing the location of the contacts of the Rostral (blue) and Caudal (dark grey) leads with respect to the midline (in dashed red). **(c)** Location of the motoneurons of arm and hand muscles in the human spinal cord in relation to spinal segments (light yellow) and vertebrae (grey). We estimated the rostro-caudal position of motoneuron pools from Schirmer 2011. **(d)** Graphical representation of muscle activation obtained by stimulating through selected contacts (labelled in red on the left of each human figurine). Each human figurine represents the front view (left half) and back view (right half) of arm muscles (See also **Extended Data Figure 3**). Each muscle is colored with a color scale (on the left) representing the normalized peak-to-peak amplitude of EMG reflex responses obtained during 1 Hz stimulation at the stimulation amplitude indicated on the left. Peak-to-peak values for each muscle are normalized to the maximum value obtained for that muscle across all contacts and all current amplitudes. On the left, MRI of each of the participant is shown with segmented lesion in red.

Our study was designed to quantify the immediate, assistive effects of SCS on post-stroke motor deficits, including muscle weakness, impaired dexterity of arm and finger movements, intrusion of aberrant flexor synergies and spasticity. Based on previous work in SCI, we expected any benefit to reverse once SCS was turned off^8,24–26,28^, and we did not incorporate activity-based training exercises into the protocol. Therefore, we focused on measuring immediate improvements attributable to the direct effects of SCS in facilitating motor function in the arm and hand, rather than long-term recovery. Testing began four days after implantation of the SCS leads and continued for four weeks, during which the participants participated to assessments 5 times per week. After 29 days the percutaneous leads were removed. We evaluated function with and without stimulation that we delivered continuously through a custom-built microcontroller-based system connected via percutaneous access to the SCS leads participants (**Extended Data Figure 2**).

### SCS achieved segment-level specific muscle activation in the cervical spinal cord

Clinical SCS leads are intended to be placed along the midline to broadly stimulate dorsal columns and block pain signals^39^. However, we have shown previously in monkeys and humans that selective recruitment or primary afferent fibers in the cervical dorsal rootlets can be achieved by positioning the clinical SCS leads laterally, near the dorsal root entry zone^8,10,38^. These primary afferents innervate motoneuron pools according to a well-defined rostro-caudal somatotopy^40^, and we predicted that stimulating specific nerve roots would lead to excitation of the corresponding motoneurons. For example, in humans, motoneurons from the C4 spinal segment innervate proximal muscles such as the trapezius and deltoids; while segments C8 and T1 innervate wrist extensors and hand muscles^40^ (**Figure 1c**). Consequently, we hypothesized that selective stimulation of rostral roots would facilitate muscle activation in the upper arm, while stimulation of the caudal roots would target distal muscles including the forearm and hand^10^. Therefore, we designed a surgical approach to implant two linear electrodes mediolaterally spanning the dorsal roots C4 to T1 (**Figure 1b**). During implantation, we guided surgical placement with neurophysiological intraoperative monitoring^26^ and verified that reflex-mediated muscle responses could be obtained reliably across all muscles of the arm and hand. Intra-operative data showed that SCS followed a clear rostro-caudal segmental specificity in both participants (**Figure 1d** and **Extended Data Figure 3**) Monopolar stimulation of rostral contacts induced activity in the deltoids and trapezius while caudal contacts recruited intrinsic hand muscles (**Figure 1f** and **h** and **Extended Data Figure 3**). To verify that stimulation responses resulted from afferent-mediated recruitment of the motoneurons, and not by directly recruiting ventral roots, we stimulated through the same contact at different frequencies (1.1, 2, 5, 10, and 20 Hz). Indeed, reflex mediated responses are well known to show frequency-dependent suppression phenomena^10,18^. The peak-to-peak amplitude of evoked muscle activity was reduced significantly in a frequency dependent manner confirming that motor neuron activation was occurring trans-synaptically (**Extended Data Figure 4**). Repeated X-rays showed minimal rostro-caudal displacement of the leads from the implant (**Extended Data Figure 1**) which did not affect functional performances and we utilized the same contact configurations from week 2 (when they were finalized) to week 4 (**Extended Data Figure 5**). In summary, we showed that accurate placement of clinical leads over the dorsolateral cervical spinal cord produces selective muscle activation according to well-described myotomal maps and that stimulation activates motor activity through sensory afferents in the dorsal roots.

### Arm and hand strength immediately improved upon activation of SCS

To determine whether SCS would lead to an increase in strength, we asked participants to apply their maximum force during isometric flexion and extension of single arm joints. Forces were applied to a robotic platform which measured joint torque (HUMAC NORM) (**Figure 2g**). We compared torques produced with and without continuous SCS targeting muscles of the tested joint (**Figure 2**). We found that SCS01 consistently improved shoulder, elbow and wrist strength (**Figure 2** Error! Reference source not found.**a**,**c**,**d**,**e**,**f**). Mean torques more than doubled for elbow (Day 9: 9.8 vs 22.0 Nm; Day 23: 11.6 vs 24.6 Nm). As an example of the functional relevance of these improvements, we show a video in which SCS01 can raise her arm above her head during SCS (**Video 1**). In SCS01, we tested multiple stimulation frequencies (20, 40 and 60Hz) during elbow flexion and extension isometric tests and found that 60Hz yielded maximal torques. Although the severity of SCS02’s impairment hindered consistent assessment of all joints, she did show improvement of detectable movements, including significant improvements in elbow flexion (**Figure 2a,e**) similar to those observed in SCS01 (40% average increase). We also tested isometric grip strength using a hand-held dynamometer (**Figure 2f**). SCS led to 40% increase in SCS01 and 108% in SCS02 suggesting that SCS can potentiate both arm and hand function. This result was particularly striking for SCS02 who had near complete hand paralysis and was unable to consistently produce detectable hand grip forces without SCS. Additionally, on the first day of testing, SCS01, for the first time in >5 years since her stroke, immediately reacquired the capacity to fully and volitionally open her hand (**Video 1**). We also compared the root mean square values of EMG signals measured from the anterior deltoid, biceps, and triceps during elbow extension (SCS01) and elbow flexion (SCS02). EMG was substantially higher with stimulation than without for these muscles in both participants (>100% **Figure 2b-e**) indicating that SCS potentiates the participant’s ability to recruit muscles.

**Figure 2.**
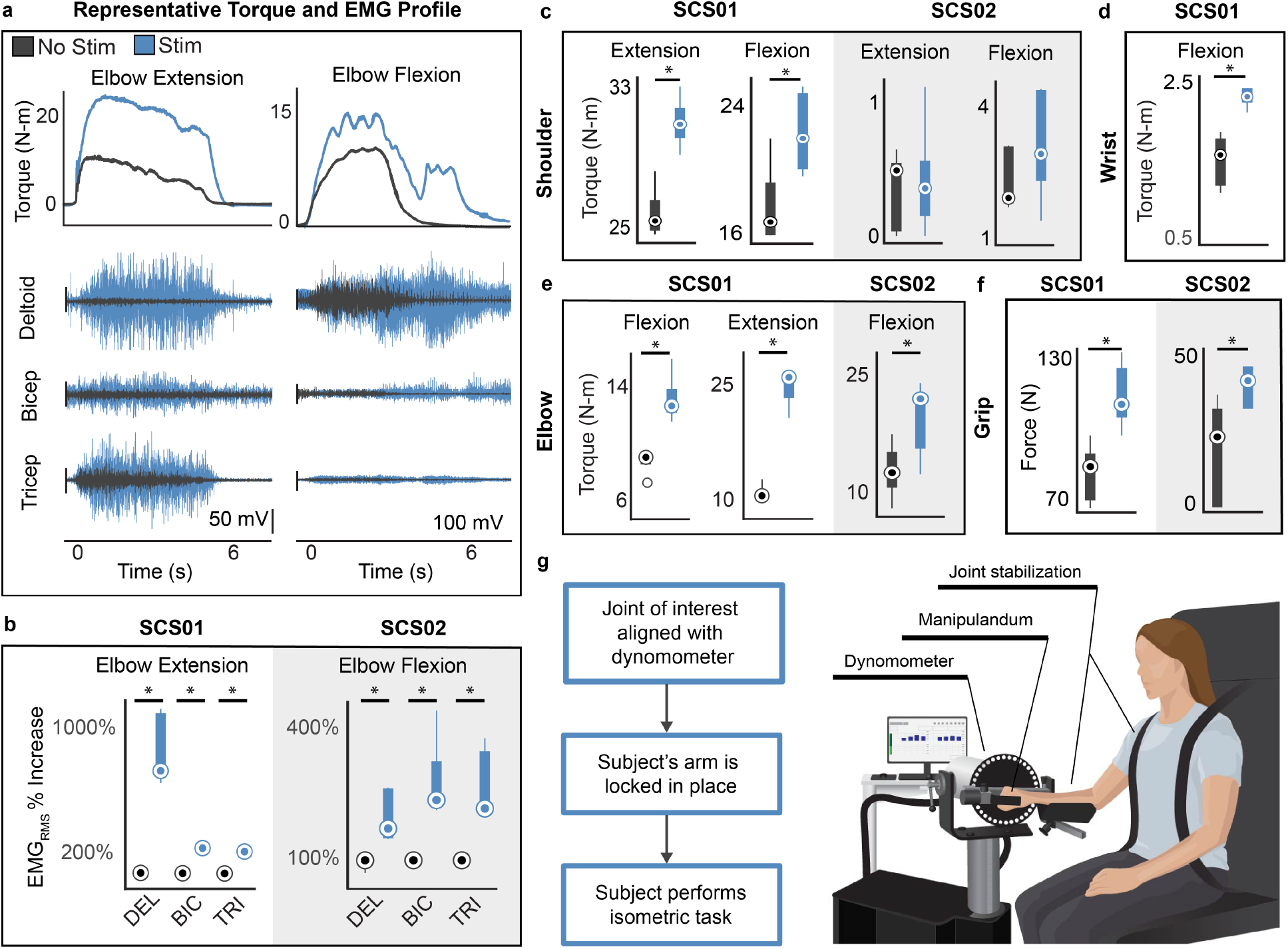
SCS immediately improves strength. **(a)** examples of single synchronized raw traces for torques and EMGs signals during isometric maximum voluntary contractions for extension (SCS01, left) and flexion (SCS02, right) of the elbow in the HUMAC NORM (see panel **g**). **(b)** quantification of the root mean square value of EMG traces with and without stimulation during isometric elbow extension (SCS01) and flexion (SCS02) **(c, d, e)** quantification of isometric torques during single joint flexion and extension for SCS01 and SCS02 at shoulder, elbow and wrist **(f)** quantification of isometric grip-strength measured with a hand-held dynamometer with and without stimulation. **(g)** schematic of the isometric torque test (wrist configuration in the example) in the HUMAC NORM. *Statistics* all quantifications are reported using box-plots. Median values are represented by circles. Inference on mean differences is performed by bootstrapping the n=5 repetitions obtained for each measurement, with n=10,000 bootstrap samples; * difference is outside the resulting 95% confidence interval.

Given that SCS results in a sensory perception known as parasthesia^38^, it was impossible to rule out placebo effects because participants where always aware that stimulation was active. To test for the effects of motivation and bias, we performed sham trials in which non-optimal stimulation was delivered without participant knowledge. In these sham trials, we selected electrodes that preferentially activated muscle groups that were antagonistic to the movement performed. SCS01 still experienced paresthesia over the shoulder and arm during stimulation and was unable to distinguish optimal from sub-optimal configurations. As expected, while even antagonist stimulation led to some increase in strength (19% (extension) and 16% (flexion) greater than no stimulation), the most significant improvements in strength occurred only when SCS was optimized for the intended movement (agonistic stimulation; 82% (extension) and 25% (flexion) greater than no stimulation) (**Extended Data Figure 7**). In summary we showed that SCS led to immediate and substantial improvements in strength and muscle activity of the arm and hand.

### SCS improved arm motor control during planar reaching

In addition to strength, we evaluated the benefit of SCS on arm dexterity and muscle synergies. For this, both participants performed planar reach and pull tasks using a robotic platform (KINARM) that supported the weight of their arm (**Figure 3a**). Importantly, these reaching tasks were performed in the horizontal plane to dissociate the effects of shoulder weakness and compensatory movements from the capacity of participants to extend their arm towards a target^41^.

**Figure 3.**
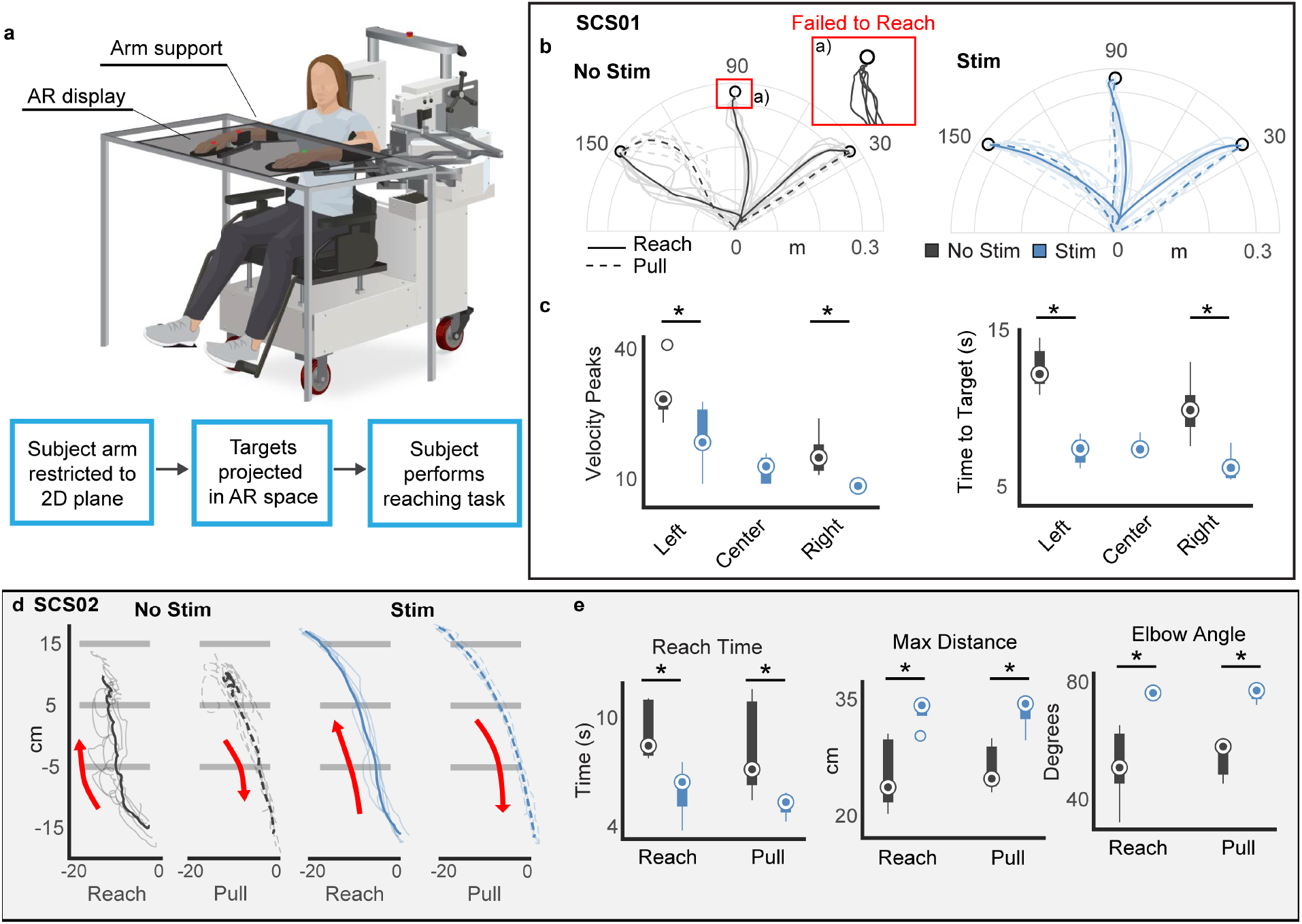
SCS immediately improves arm kinematics. **(a)** schematic of the experimental set-up for planar reach out tasks using the KINARM. **(b)** Examples of raw endpoint trajectories for SCS01 in the reach out task. No stimulation on the left (dark grey) and during stimulation on the right (blue). Inset shows inability to reach central target with no stimulation. Solid lines are reach trajectories and dashed lines represent pull trajectories. Darker lines represent average trajectories, shaded lines represent single trajectories. **(c)** Quantification of kinematic features, movement smoothness (velocity peaks) and time to reach target in s. Center target could not be calculated for no-stim condition because SCS01 did not complete the task. **(d)** Examples of raw endpoint trajectories for SCS02 in the reach out task. SCS02 was tasked to reach beyond the third horizontal line to complete the task. Reach and pull trajectories are represented in separate plots. **(e)** Quantification of kinematic features for SCS02, Reach time (equivalent to time to target in SCS01), Maximum reached distance and elbow angle excursion (max-min) are reported for no-stim (dark grey) and stim condition (blue). *Statistics* all quantifications are reported using box-plots. Median values are represented by circles. Inference on mean differences is performed by bootstrapping the n=6 (center out) or n=5 (open-ended reaching) repetitions obtained for each measurement, with n=10,000 bootstrap samples; * difference is outside the resulting 95% confidence interval.

SCS01 was tasked with reaching towards targets positioned to maximize active elbow extension since this was particularly difficult for the participant due to the intrusion of flexor synergies. During stimulation, SCS01 was able to successfully reach all targets; whereas, without stimulation, she was never able to reach the central target because of difficulty extending her arm (**Figure 3b**). Movements to targets that she could consistently reach with and without stimulation, were significantly faster and smoother with stimulation on (**Figure 3c**; 34% (left target) and 47% (right target)). Similarly, speed (**Figure 3c**) trajectory variability and max distance reached, were all improved with stimulation compared to controls (**Extended Data Figure 6b**). These results indicate that during stimulation SCS01 was able to perform smoother, faster, and more accurate movements as well as reach targets that she could not reach without stimulation.

Due to the severity of SCS02’s impairment, she performed a simpler task in which she was instructed to reach the furthest of three horizontal lines spaced at 10 cm intervals (**Figure 3d)**. Without stimulation, SCS02 was never able to reach the farthest line, but with stimulation on she was able to reach it on every trial due to the facilitation of elbow extension. This was reflected in the elbow excursion angle, which increased 23 degrees with stimulation (**Figure 3e**). The maximum distance reached was 7.8 cm greater and total movement time was 37% faster with stimulation, **Figure 3e**. Like SCS01, her movements also became smoother during stimulation (20% fewer velocity peaks **Extended Data Figure 6c**), and her trajectory variance and total path length also significantly improved (**Extended Data Figure 6c**). Arm extension kinematics and elbow angle were strongly modulated by stimulation frequency in SCS02 showing peak performances at 100Hz (**Extended Data Figure 6a**).

We hypothesized that the improvements in reaching function were attributable to facilitation of elbow muscle activity and changes in flexion and extension synergies. To test this, we inspected EMG activities and extracted muscle flexor and extensor synergies associated with the extension and flexion movement phases using dimensionality reduction (**Extend Data Figure 8**, see methods). Without stimulation, muscle activity was very low at the elbow muscles and very high at the shoulder muscles, likely indicating a compensatory strategy dominated by shoulder muscles and allowing the elbow to extend passively during the reach. This was reflected in the strength of the components of each synergy that showed a greater contribution of shoulder muscles in both participants. Instead, with stimulation, the contribution of elbow muscles increased significantly and became dominant in both synergies which corroborates our hypothesis and demonstrates a reduction of compensatory shoulder movements.

To test if stimulation specificity was necessary for optimal motor control, we performed a sham-controlled task in which non-optimal stimulation was delivered without participant knowledge in the center out task. **Extended Data Figure 7** shows the dramatic impact of incorrect stimulation configuration on SCS02’s task performance. Specifically, during sham-stimulation, arm kinematics suffered dramatically and her performance worsened, even compared to her ability with stimulation off, significantly affecting her ability to reach designated targets. Instead with optimal stimulation she reached all targets 100% of the times.

In summary, we showed that SCS targeting dorsal roots at specific cervical segments improved dexterity and enabled participants with stroke to perform smooth and effective arm movements enabling full elbow extension improving elbow extension and flexion synergies and reducing compensatory shoulder activity.

### Functional benefits of SCS

Finally, we sought to determine whether these improvements in strength and control translated to functional and daily life activities (**Figure 4**). For this, we personalized tasks according to impairment level. We first evaluated the ability of SCS01 to perform 3D reaching movements. We asked SCS01 to reach as fast as she could towards 6 targets placed on two different horizontal planes that required both planar and upward reaching movements against gravity. SCS enabled her to reach all targets faster, approximately reducing in half the total task-completion time (**Figure4f**). We also asked SCS01 to perform a classic manipulation task: the box and blocks task, in which she was instructed to move small cubic objects from one side of a box to the other by grasping and lifting them over a barrier. During stimulation she was able to perform this task nearly twice as fast (**Figure 4e, Video 4**). Finally, we increased the complexity of the tasks by presenting daily living tasks that required high-skill and dexterity such as drawing a spiral, reaching for and lifting a soup can, eating with a fork, and opening a lock. SCS increased her overall dexterity, allowing her to produce smoother and more consistent drawings (**Figure 4a**). SCS enabled simultaneous reaching, forearm supination and grasp allowing SCS01 to reach, grasp and lift a soup can. Without stimulation, forearm pronation and supination were not possible (**Video 2**). This improvement of fine motor skills even enabled her to grasp a key and open a lock, whereas without stimulation she was unable to hold the key or lock at all (**Figure 4c**). Finally, with stimulation, SCS01 was able to feed herself using her affected hand, with full independence; a task that she had been unable to perform for 9 years (**Video 3**).

**Figure 4.**
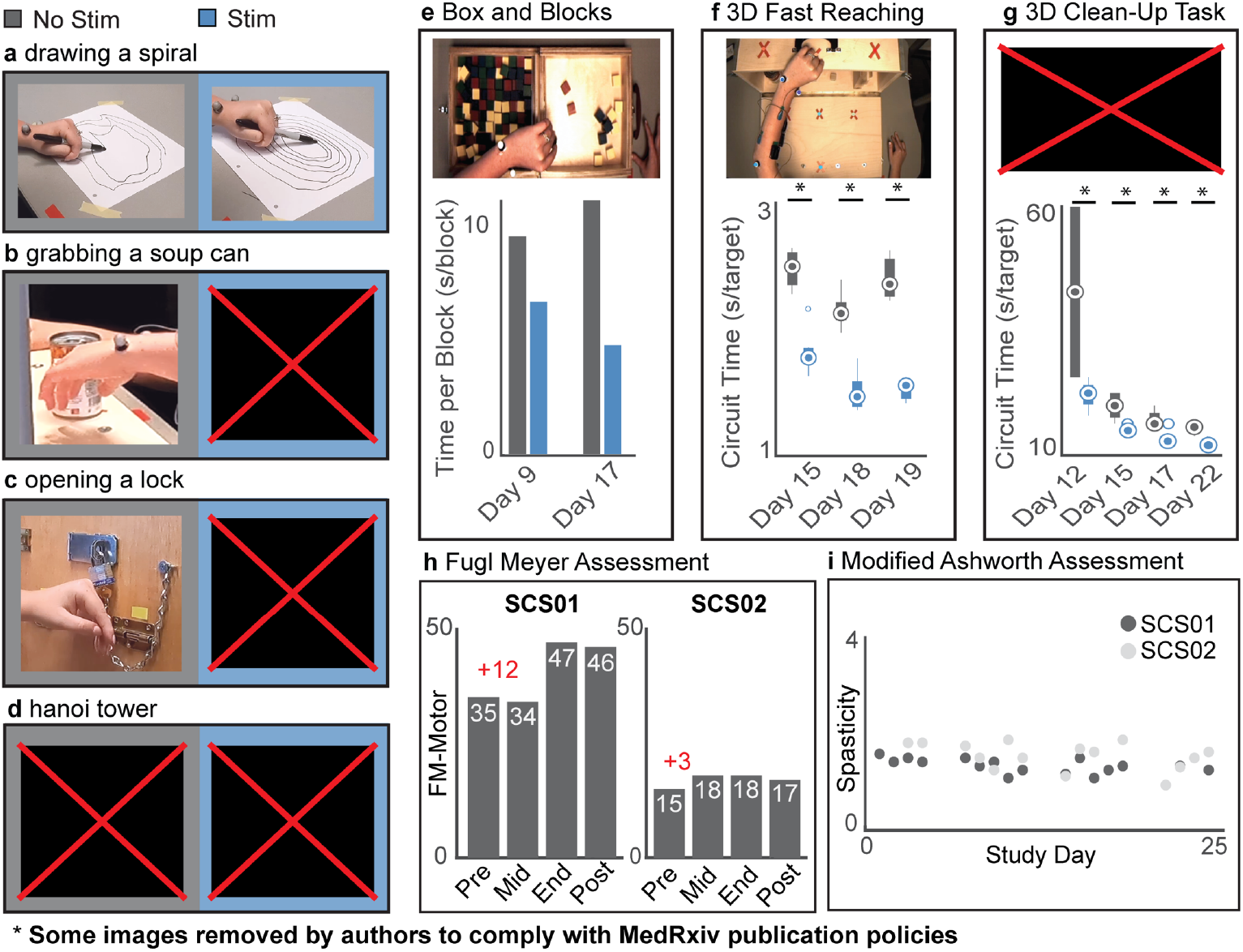
SCS improves function. ***(a***,***b***,***c)*** frame captures from videos showing improved functional abilities of different simulated activities of daily living: drawing a spiral, reaching and grasping a soup can, opening a lock for SCS01. Left no stimulation, right with stimulation. **(d)** picture report frames from video of SCS02 performing a modified “Hanoi tower” task in which she was task to move a hollow cylinder from a base pole to another. Left no stimulation, right with stimulation. **(e**,**f)** representative pictures and quantification of task performances for SCS01 box and blocks and 3D fast reaching task performed on multiple days. **(g)** picture of the 3D reaching task using the Armeo Power for SCS02 and relative task performance on multiple days. **(h)** Fugl-Meyer assessment ad different time points for SCS01 and SCS02 including 4-weeks post-study. **(i)** normalized spasticity level obtained by averaging Modified Ashworth Score at each joint for SCS01 (dark grey) and SCS02 (light grey). *Statistics* all quantifications are reported using box-plots. Median values are represented by circles. Inference on mean differences is performed by bootstrapping the n=5 repetitions obtained for each measurement, with n=10,000bootstrap samples; * difference is outside the resulting 95% confidence interval. **Some images removed by authors to comply with MedRxiv publishing policies. Please contact the corresponding author for more information**

Since SCS02 was unable to sustain the weight of her arm against gravity, we questioned whether the assistive effects of SCS would improve motor performance in a typical rehabilitation robot. Therefore, we tested the participant’s performance in a clinically-approved powered-exoskeleton (Hocoma Armeo Power) that allows for titrated assistance. While in the robot, she performed tasks that required 3D reaching movements similar to those performed by SCS01 (**Figure 4g**). With stimulation, SCS02 was significantly more efficient at the task and managed to consistently reach towards more targets than without stimulation across three sessions (**Figure 4g)**. We then asked her to perform a skilled motor task where she had to remove a hollow cylinder from a wooden dowel and slip it over another. With SCS she was not only able to grasp and lift the metal cylinder, but also to place it on the adjacent dowel without any weight support (**Figure 4d** and **Video 5**). These combined results show that the assistive effects of SCS can facilitate large improvements on functional tasks and activities of daily living.

### Spasticity, tolerability, and unexpected lasting effects on motor control

To ensure that SCS did not exacerbate spasticity, we measured the Modified Ashworth Scale on each day of testing. Over the course of four weeks, we found that SCS did not lead to any worsening in spasticity (**Figure 4i** and **Extended Data Table 2**). In addition, the two participants did not report increased rigidity nor painful sensations during SCS. In fact, both patients described the stimulation as a “feeling of power in the arms” or a feeling of “being able to control my arm as if I know what I should do to move it”. Since this pilot was designed to study the assistive rather than the therapeutic effects of SCS, participants did not receive concomitant physical therapy over the four weeks. We thus did not expect to observe sustained improvements when SCS was turned off. Nevertheless, when we compared the participants’ pre-and post-study Fugl-Meyer scores, SCS01 improved from 35 points at enrollment to 47 points, and SCS02 from 15 points to 18 points. These scores were retained at 4 weeks after the end of the SCS period (**Figure 4h** and **Extended Data Table 1**).

## DISCUSSION

In this study we demonstrate for the first time that continuous SCS targeting cervical dorsal roots immediately improves upper limb strength, motor control, and function in humans with moderate to severe post-stroke hemiparesis. This assistive effect is lost when SCS is turned off. However, both participants showed some lasting improvements in motor function by week 4 of the study that were retained even without stimulation.

### Spinal cord stimulation improves cortico-spinal control in chronic stroke

Although SCS for spinal cord injury (SCI) has recently generated considerable excitement^25–27^, anecdotal reports of the beneficial motor effects of SCS in people with SCI, multiple sclerosis, cerebral palsy and even stroke date back more than 40 years^42,43^. Unfortunately, a lack of understanding of the mechanisms of SCS led to considerable variability in implant location, which affected the size and consistency of the observed effects. We now know that SCS engages spinal motoneurons via recruitment of primary sensory afferents, providing pre-synaptic excitatory input to motoneurons and other spinal interneurons directly connected to these afferents^6,10,21,22,33^. Thus, it could be hypothesized that by raising the membrane potential of neurons in spinal circuits, SCS could increase responsiveness to residual cortical inputs and immediately improve voluntary motor control^10,23,34^. We define this as the “assistive effect” of SCS. By implanting epidural electrodes over the lateral aspect of the cervical spinal cord, we focused this assistive effect on the arm and hand motor pools most needed for each participant^8,10^. While our approach enabled a high degree of personalization of stimulation patterns, we argue that the large effect sizes we measured were possible because, unlike in SCI, the stroke lesion spares cervical spinal circuits and usually some supraspinal pathways. Indeed, studies of cervical SCS in SCI have not shown the magnitude of immediate assistive effects at the arm and hand as those that we report here post-stroke^11,44^.

### Relevance for clinical therapy of post-stroke hemiparesis

The selective targeting of specific dorsal roots allowed us to use simple yet personalized continuous stimulation protocols. Our surgical approach made this personalization straightforward by leveraging clear somatotopic organization of the cervical spinal cord and the final configurations of stimulating electrodes were selected during the first 2 weeks of the study. We believe that the simplicity and robustness of our protocol could be translated readily to the clinic. Indeed, the implantation and programming procedures are similar to those performed routinely in the application of SCS for refractory pain, which accounts for tens of thousands of patients every year^45^. While non-invasive alternatives to SCS are being investigated in SCI^44^ our results in stroke depended on fine tuning of stimulation parameters at particular contacts, which would not be possible with the limited specificity of transcutaneous SCS^46^.

Here we placed an emphasis on quantifying the immediate assistive effects of SCS. In contrast, almost all rehabilitative stroke studies concern recovery after the intervention is over^1,47,48^ which makes comparison with existing literature difficult. That said, although our two participants received no concomitant training over the four-week study, we observed some persistent motor recovery that was particularly large in SCS01. We believe this is a promising indication that SCS may promote a true post-stroke restorative effect that goes beyond assistance. This restorative avenue is especially exciting considering the advent of new and more effective impairment-focused behavioral interventions for stroke^49–51^ that could be combined with SCS into a realistic yet effective therapy for post-stroke hemiparesis.

## Data Availability

All software and anonymized data will be available upon reasonable request to the corresponding author.

## AKNOWLEDGEMENTS

We thank Jemère Ruby for the design of figure elements. We wish to thank Tyler Simpson for the engineering support provided. The study was executed through the support of internal funding from the Department of Neurological Surgery at the University of Pittsburgh to MC. Additional funding was provided by the Department of Mechanical Engineering and the Neuroscience Institute at Carnegie Mellon University to DW and the Department of Physical Medicine and Rehabilitation at the University of Pittsburgh to EP.

## AUTHORS CONTRIBUTIONS

MC, DW and EP conceived the study. MC, DW and EP secured funding. MC, EP, JK and DW designed the experiments and assessments. MP, DW, NV and ES designed and implemented the stimulation control system, hardware and software framework for the experiments. EC and EP designed and implemented KINARM motor tasks. SE and EP designed and performed MRI-based analyses. AB and GW implemented patient recruitment, eligibility and monitoring and coordinated study management. MC, LF and PG designed the neurosurgical approach. PG, DF and RF implemented and evaluated neurosurgical procedures and participants’ clinical management protocols. JB, MP and MC designed and performed intraoperative monitoring protocols. MP, NV, ES, EC, MC, EP, SR, BB, AB and JK performed the experiments. JG designed statistical data analyses. MP, NV, ES, EC and SR analyzed the data. MP, ES, EC and NV created the figures. MC, MP, EP, DW, ES and NV wrote the paper and all authors contributed to its editing.

---

**Extended Data Figure 1.**
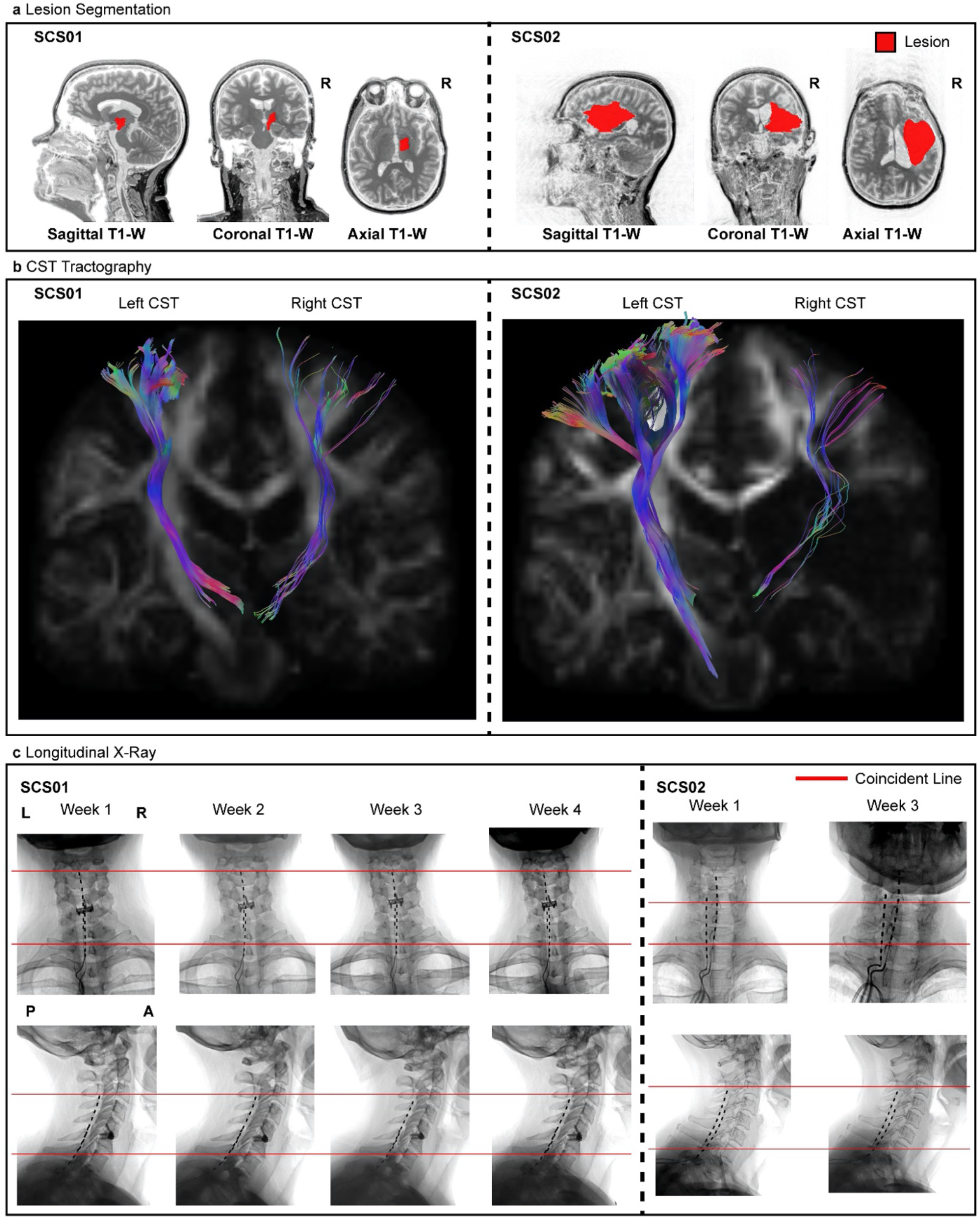
Lesion characterization and Lead position over time. **(a)** sagittal, coronal, and axial T1-weighted MRI 2D projections for SCS01 and SCS02. The segmented lesion is shown in red for both participant. R indicates the Right hemisphere. **(b)** High-definition fiber tracking of the corticospinal tract (CST) for SCS01 and SCS2. Colored fibers represent estimated CTS axons from the affected (right) and unaffected (left) hemisphere. Significant reduction in number of tracked fibers in the right hemisphere is clear in both participants in consequence of the stroke. **(c)** Repeated X-rays for SCS01 (left) and SCS02 (right) showing the position of the spinal leads. The red lines mark the same anatomical location across the X-rays to facilitate interpretation. Minimal displacement occured after initial implantation.

**Extended Data Figure 2.**
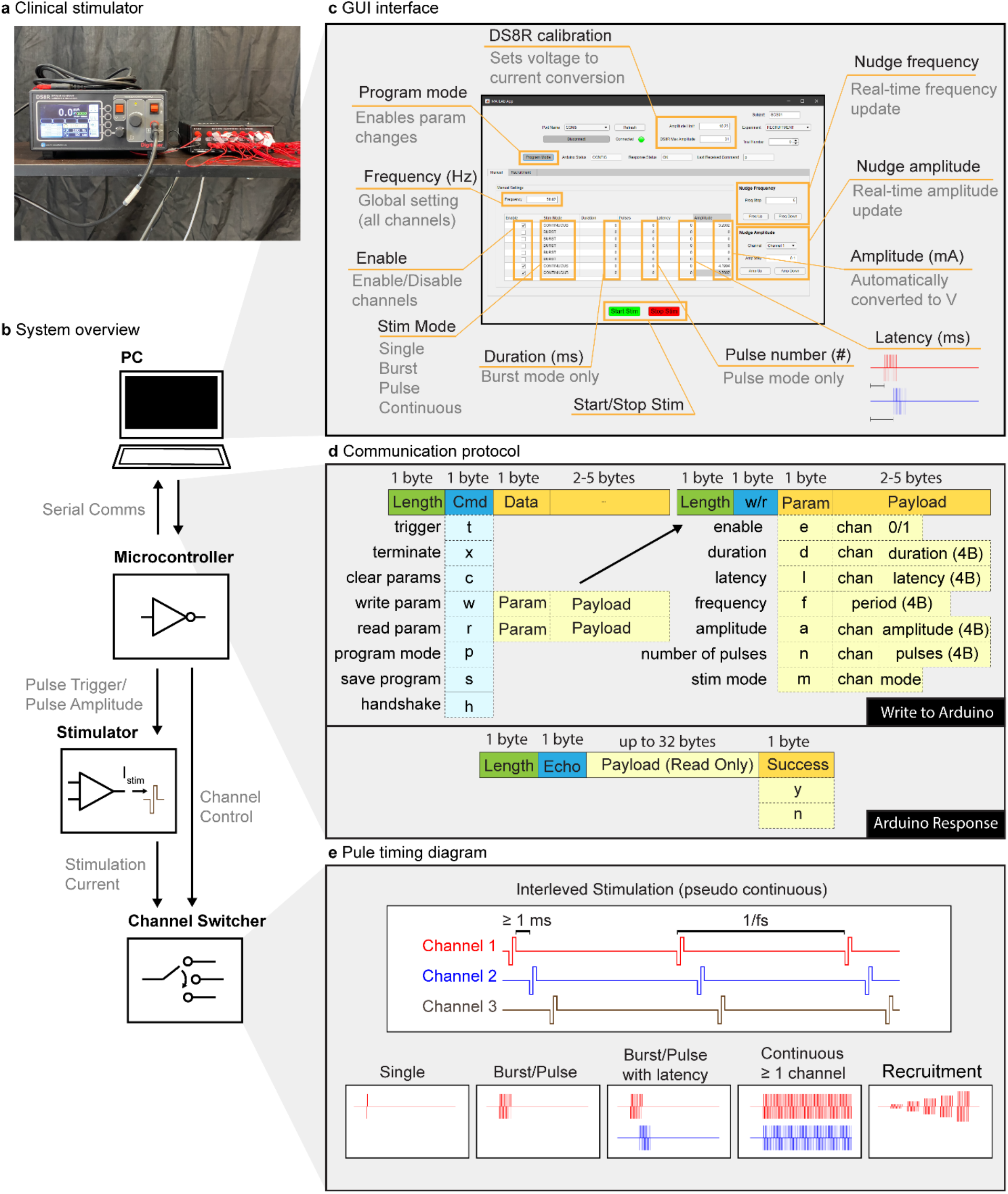
SCS parameters set using a custom-built controller. **(a)** An image of the stimulator (DS8R, left) and 1-to-8 channel multiplexer (D188, right) used to deliver stimulation pulses. **(b)** An overview of the control scheme used to deliver patterns of stimulation. A PC running a **(c)** MATLAB based GUI communicated with a microcontroller using a custom **(d)** communication protocol over a virtual serial port. The microcontroller’s firmware delivered pulse triggers and amplitude control signals to the stimulator as well as an 8 bit parallel channel selection signal to the multiplexer in order to control pulse timing, amplitude, and output channel. Current was delivered from the stimulator through the multiplexer and ultimately to the selected electrode on the implanted spinal array. **(c)** The GUI interface allowed for configuring all stimulation parameters including active channels, stimulation frequency, pulse train duration (or continuous), pulse train latency, and stimulation amplitude for each active channel. Once configured, stimulation was initiated or terminated via the software interface. The software also allowed for rapid changes in either global stimulation frequency (nudge frequency) or channel amplitude (nudge amplitude). **(d)** A custom command protocol layer was developed on top of a UART serial interface to enable communication between the GUI and microcontroller. Each packet from the master (PC) to the slave (microcontroller) comprised a 1 byte packet length, 1 byte command, and 0-6 bytes of payload. A payload comprised a 1 byte parameter (to be read or written), a 1 byte channel number (when appropriate), and the value to be written (when ‘write’ command was used). Microcontroller response packets comprised a 1 byte packet length, 1 byte command echo, 0-32 bytes of payload (used to return parameter values during ‘read’ command), and a 1 byte success flag. **(e)** The microcontroller firmware allowed for pseudo-synchonous stimulation across multiple channels by interleaving pulses on all active channels. A delay of at least 1 ms between each pulse allowed enough time for the multiplexer to fully switch channels. The same pattern of pulses was delivered every period as defined by the stimulation frequency. Each channel could also be configured to deliver a single pulse, a pulse train with finite duration and/or latency, continuous stimulation, or a ‘recruitment curve’ in which the amplitude was gradually increased for successive pulse trains of specified length.

**Extended Data Figure 3.**
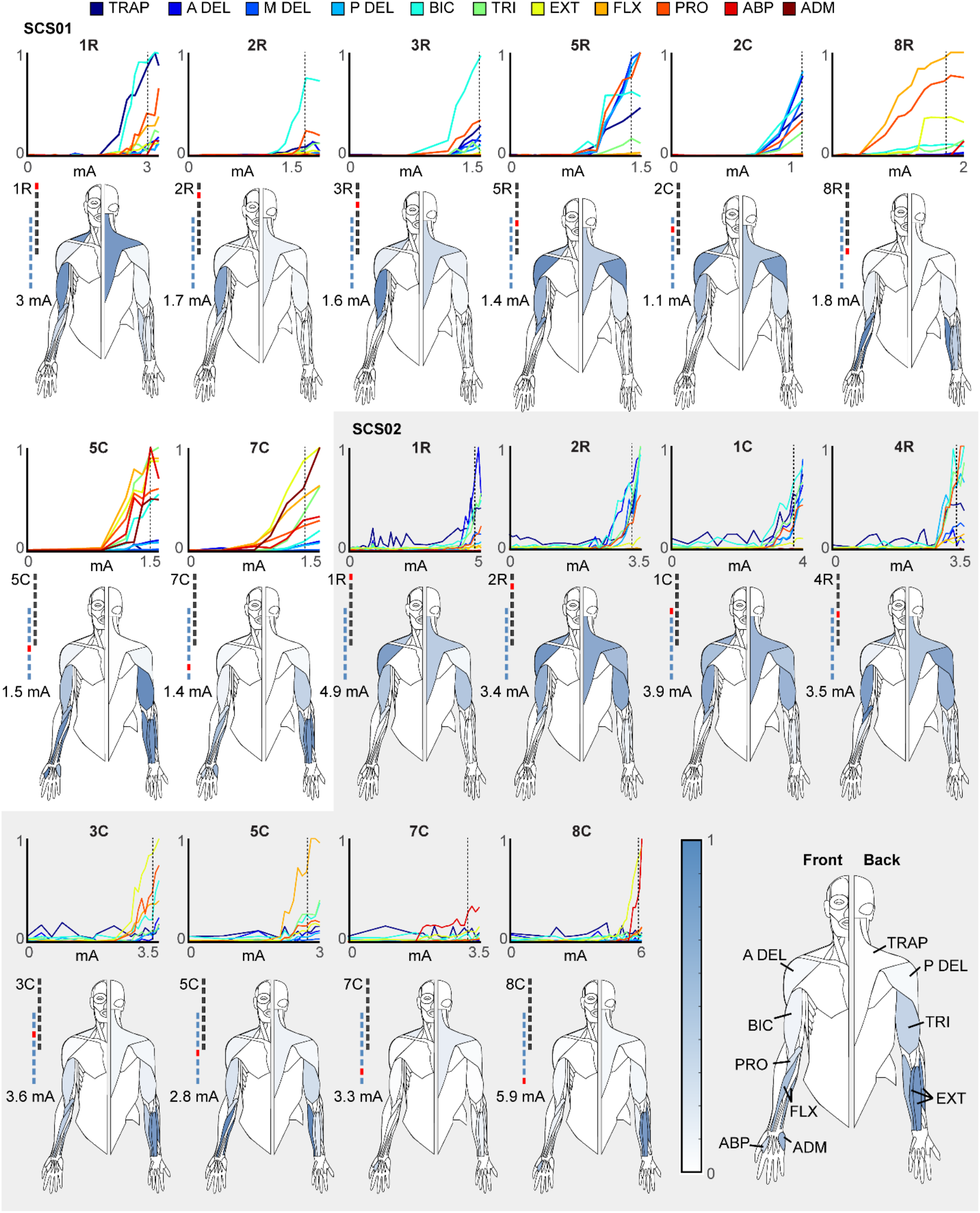
Muscle recruitment curves. In each panel we show the recruitment curves obtained with stimulation at 1 Hz at increasing current amplitude for 11 arm and hand muscles: TRAP: trapezius, A, P, M DEL: anterior, posterior and medial deltoid respectively, BIC: biceps, TRI: triceps, EXT: Extensor carpi, FLX: flexor carpi, PRO: pronator teres, ABP: abductor pollicis and ADM: abductor digiti minimi. Below each set of recruitment curves we report the graphical representation of the muscle activation obtained at the amplitude indicated on the left of each human figurine. Interpretation of human figurines is reported in the bottom right. Each muscle is colored with a color scale (on the left) representing the normalized peak-to-peak amplitude of EMG reflex responses obtained at the stimulation amplitude indicated on the left. Peak-to-peak values for each muscle are normalized to the maximum value obtained for that muscle across all contacts and all current amplitudes.

**Extended Data Figure 4.**
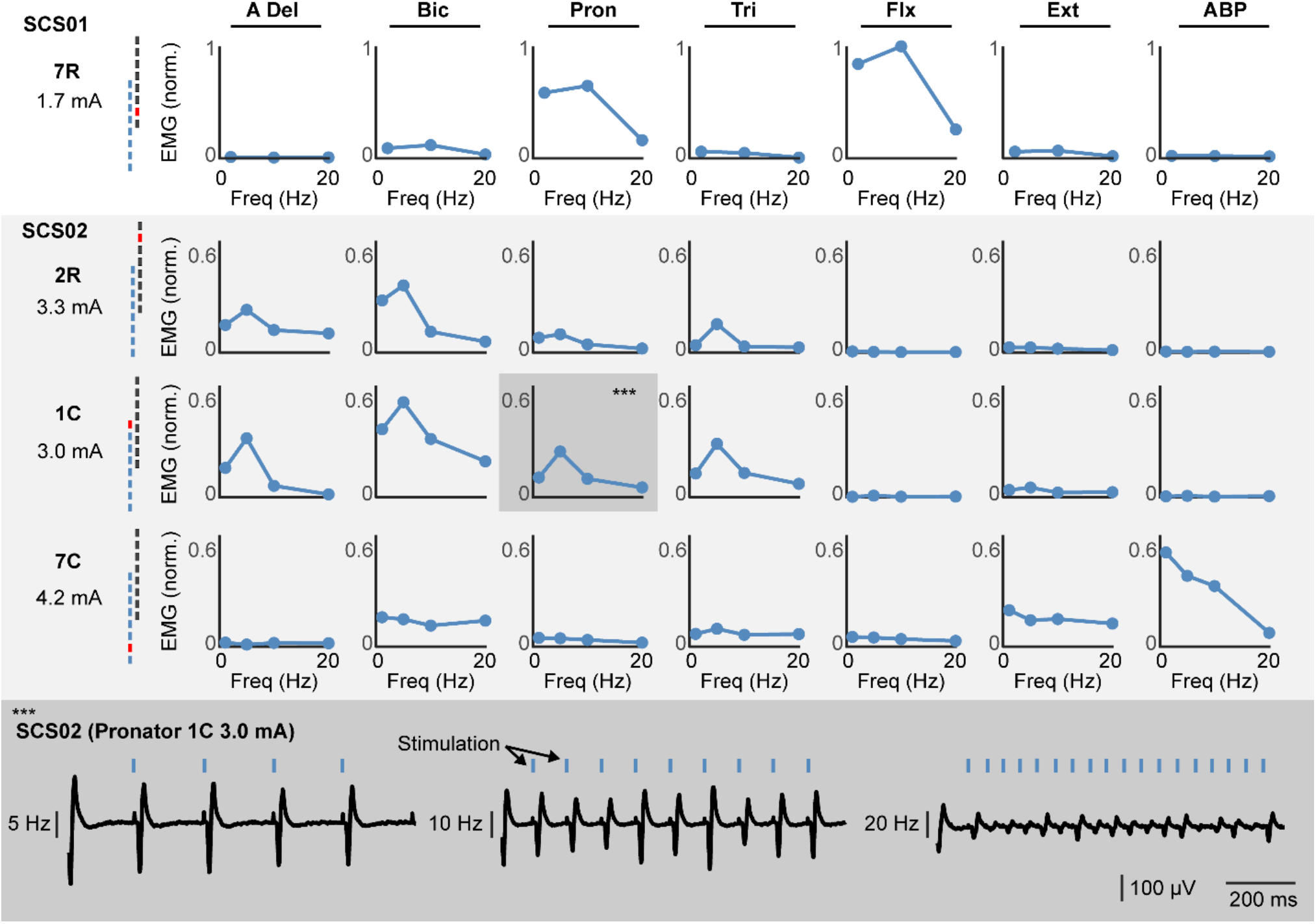
Frequency dependent suppression. To demonstrate that SCS recruits arm and hand muscles via direct activation of the primary afferents we performed stimulation at multiple frequency. The figure reports the spinal reflexes obtained when stimulating at 1, 5, 10 and 20Hz from multiple contact and multiple muscle. Each plot on the top shows the normalized peak-to-peak reflex amplitude as a function of frequency showing in the muscles that respond to the specific contact substantial frequency dependent suppression at stimulation frequencies greater than 10Hz. On the bottom, we report raw EMG traces that show the classic phenomenon. At 5Hz each pulse of stimulation corresponds to a clear evoked potential in the EMG albeit amplitude slightly diminishes at each pulse. At 10Hz, modulation of peak-to-peak amplitudes becomes more evident, at 20Hz almost complete suppression of EMG evoked responses subsequent to the first is shown. Example is taken from Pronator muscles, contact 1C, (highlighted in darker grey in the top panel).

**Extended Data Figure 5.**
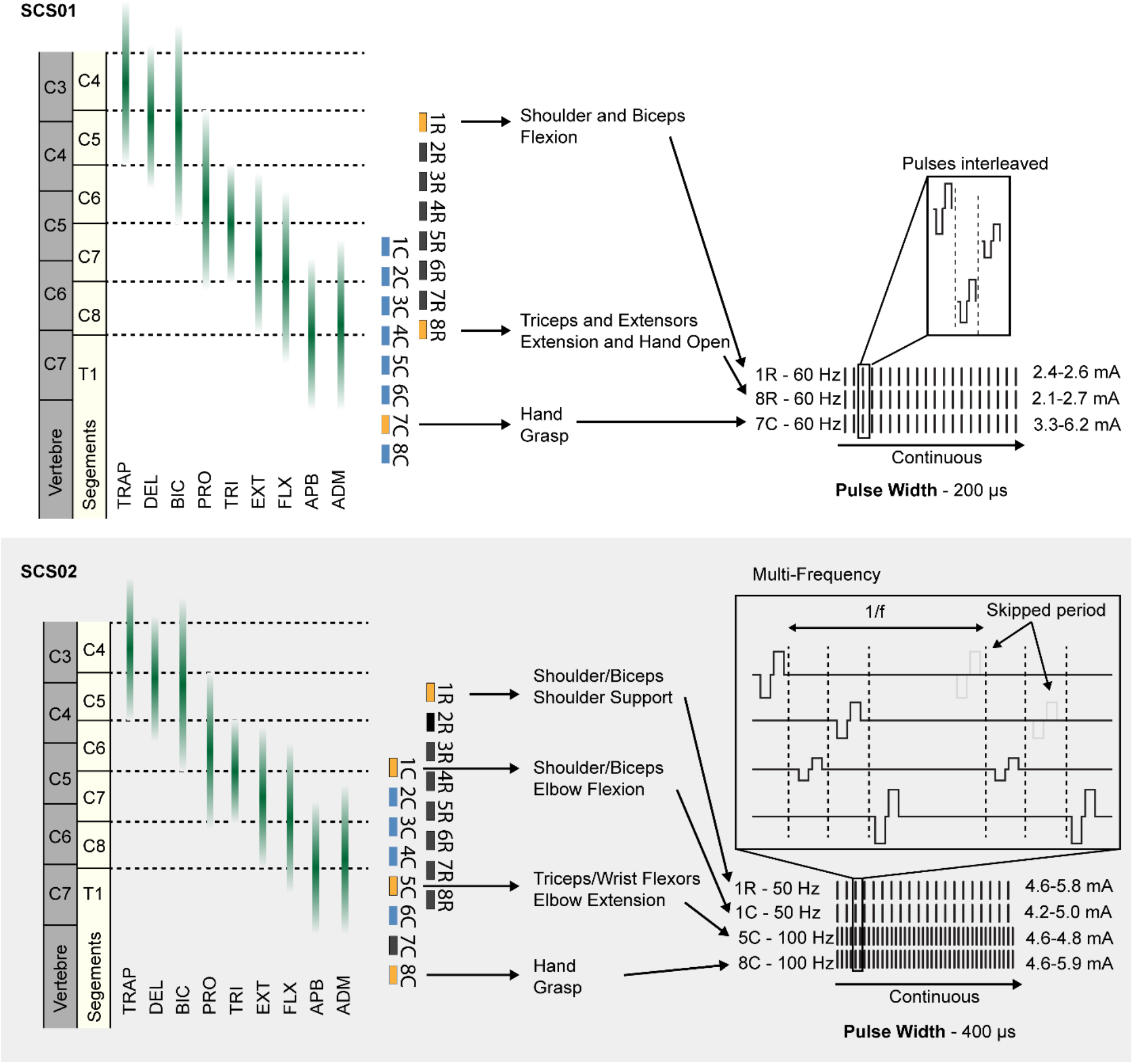
Optimized continuous stimulation protocols. Stimulation protocol used to achieve maximum assistive benefit for SCS01 **(top)** and SCS02 **(bottom). (top)** For SCS01, contacts 1R and 8R on the rostral lead and 7C on the caudal lead were simultaneously and continuously activated at a fixed 60 Hz frequency and 200 µs pulse width. These electrodes corresponded shoulders and biceps (1R); triceps, extensors, and hand opening (8R); and hand grasp (7C). Amplitudes were changed daily based on participant preference and were set to 2.4-2.6 mA (1R), 2.1-2.7 mA (8R), and 3.3-6.2 mA (7C). **(bottom)** For SCS02, contacts 1R on the rostral lead, and 1C, 5C, and 8C on the caudal lead were simultaneously and continuously stimulated. These electrodes corresponded to muscles related to shoulder support (1R); elbow flexion (1C); elbow extension and wrist flexion (5C); and hand grasp (8C). Contacts 1R and 1C were stimulated at 50 Hz while 5C and 8C were stimulated at 100 Hz all at a fixed pulse width of 400 µs. A reduced frequency was used on contacts corresponding to elbow flexion to bias the assistive benefit of stimulation toward elbow extension. Multi-frequency stimulation was achieved by skipping every other period of a 100 Hz stimulation protocol on channels stimulating at 50 Hz.

**Extended Data Figure 6.**
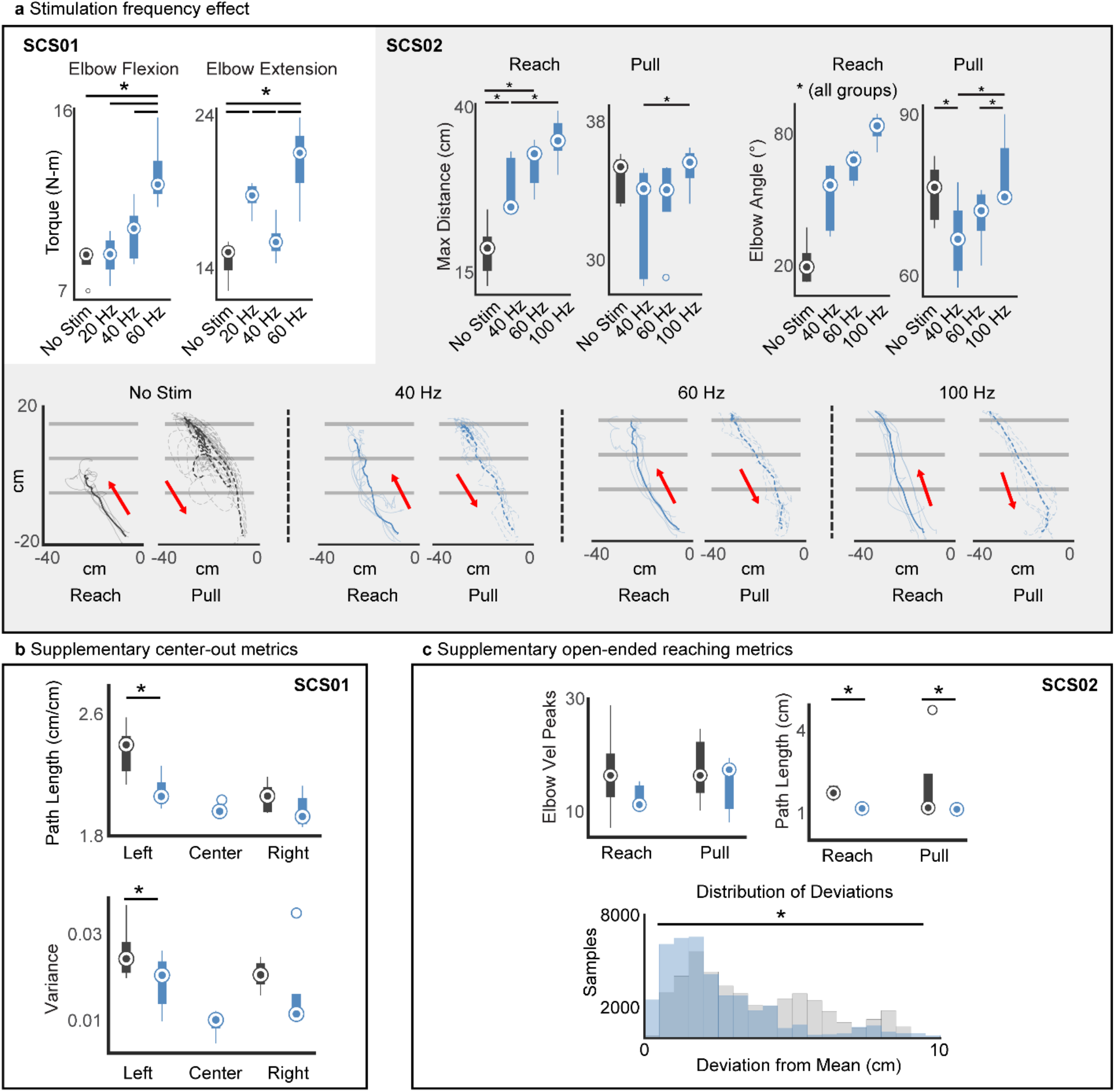
SCS improves arm kinematics supplementary metrics. **(a)** Effect of stimulation frequency shown for SCS01 and SCS02. In SCS01, quantification of isometric torques during single joint flexion and extension is shown for the elbow during no stim (dark grey), 20 Hz (blue), 40 Hz (blue), and 60 Hz (blue). In SCS02, maximum reached distance and elbow angle excursion (max-min) are reported during reach and pull of the reach-out task for no stim (dark grey), 20 Hz (blue), 40 Hz (blue), and 60 Hz (blue). Raw endpoint trajectories for SCS02 are shown in the reach out task during no stim (dark grey), 20 Hz (blue), 40 Hz (blue), and 60 Hz (blue). where SCS02 was tasked to reach beyond the third horizontal line to complete the task. Reach and pull trajectories are represented in separate plots. **(b)** Quantification of kinematic features for SCS01, path length for completed reach and pull of three targets in cm and variance of the path between trials are reported for no-stim (dark grey) and stim condition (blue). Center target could not be calculated for no-stim condition because SCS01 did not complete the task. **(c)** Quantification of kinematic features for SCS02, movement smoothness (velocity peaks) and path length in cm for reach and pull separately are reported for no-stim (dark grey) and stim condition (blue). The distribution of deviations from the mean path trajectory is shown in cm (equivalent to variance in SCS01). Inference on mean differences is performed by bootstrapping the n=5 repetitions obtained for each measurement, with n=10,000 bootstrap samples; * difference is outside the resulting 95% confidence interval.

**Extended Data Figure 7.**
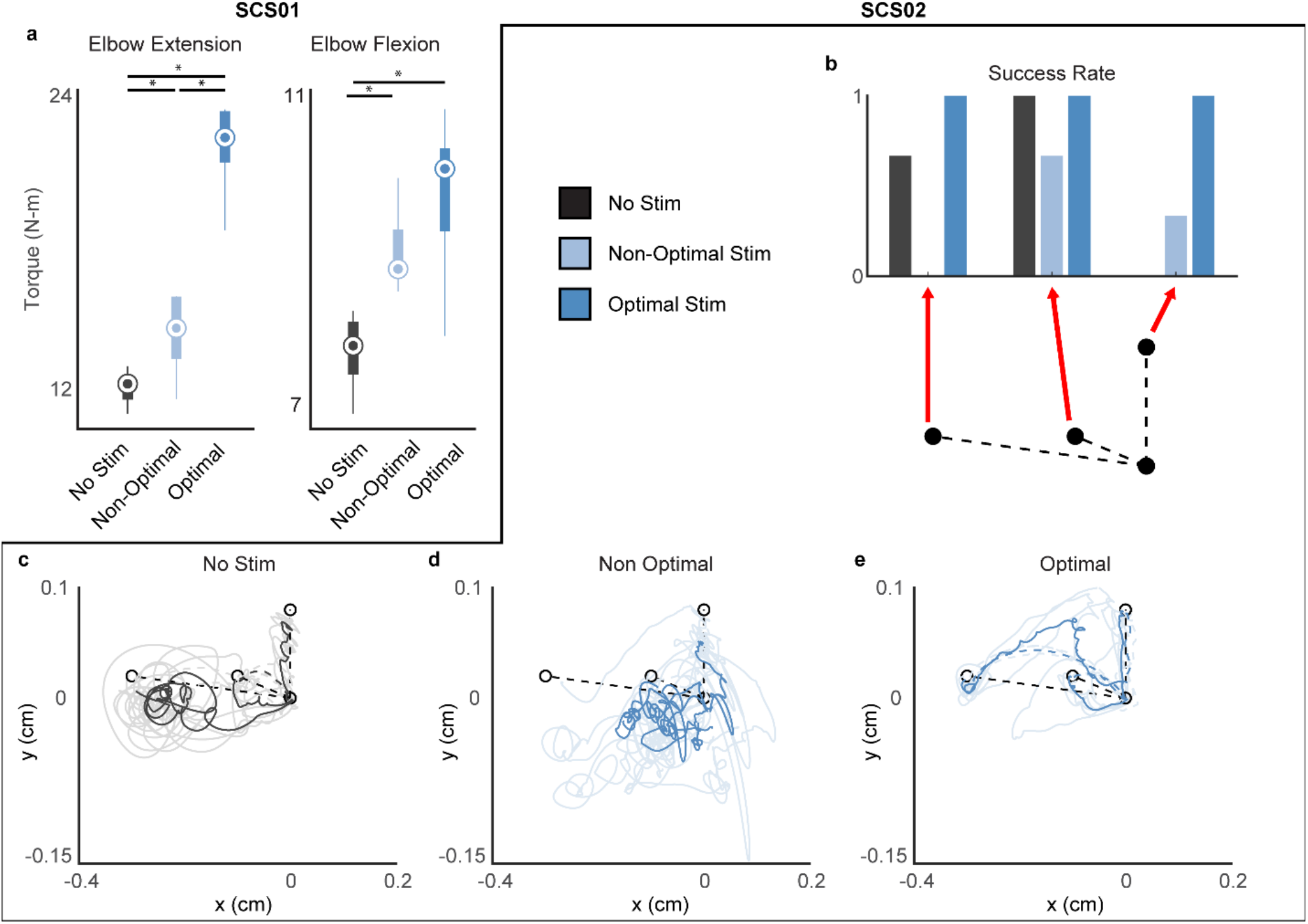
Optimized SCS leads to best improvement. **(a)** Quantification of isometric torques during single joint flexion and extension of the elbow during no stim (dark grey), non-optimal stim (light blue), and optimal stim (blue) for SCS01. **(b)** Quantification of performance for three targets of the center-out task during no stim (dark grey), non-optimal stim (light blue), and optimal stim (blue) normalized from 0 (SCS02 never reached target) and 1 (SCS02 reached target in all trials). n=3 **(c-e)** Raw endpoint trajectories by SCS02 for three targets of the center-out task during no stim (dark grey), non-optimal stim (light blue), and optimal stim (blue). Inference on mean differences for **(a)** were performed by bootstrapping the n=5 repetitions obtained for each measurement, with n=10,000 bootstrap samples; * difference is outside the resulting 95% confidence interval.

**Extended Data Figure 8.**
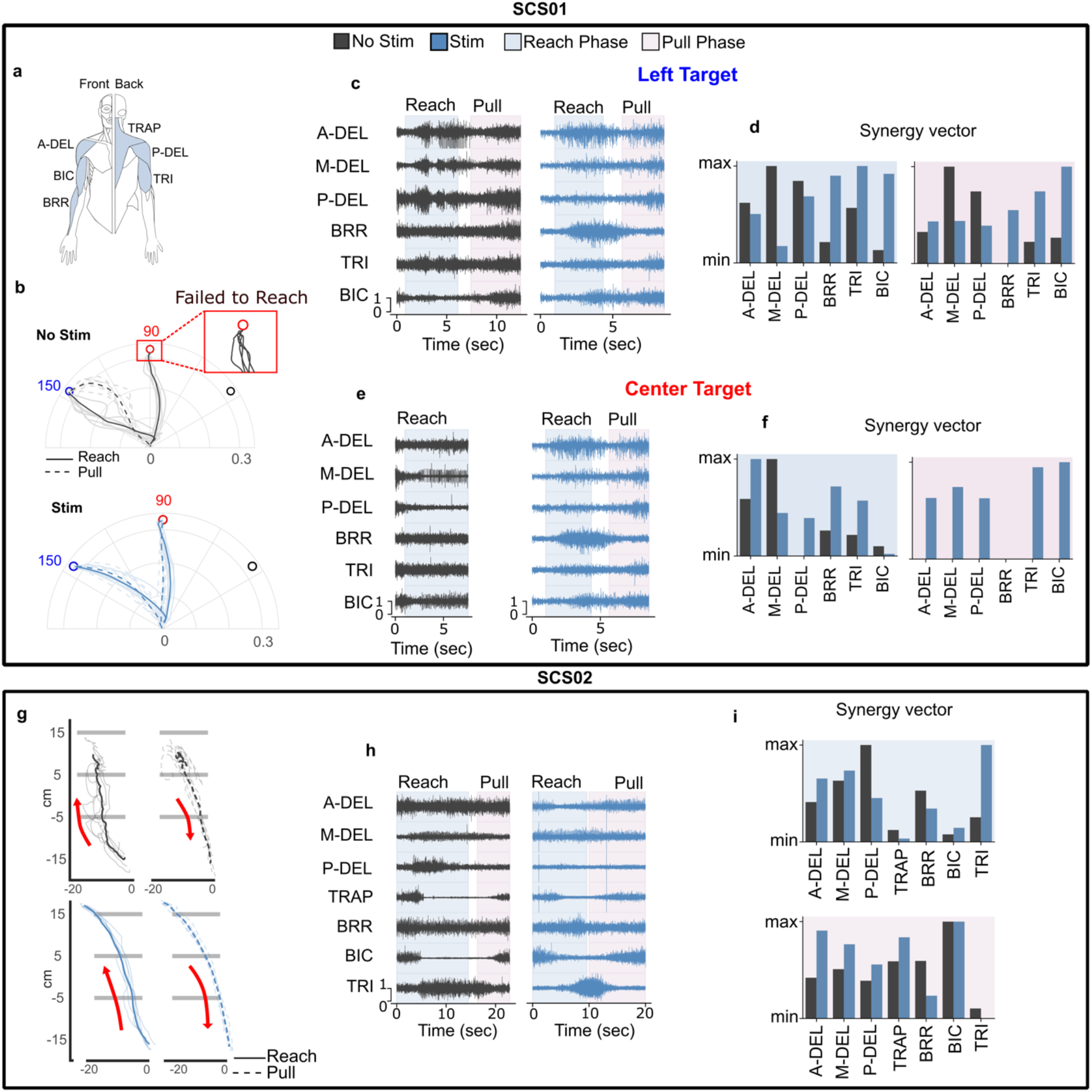
Muscle activation pattern during planar movement. **a)** Muscle label abbreviation used in the figure **(b)** Kinematic trajectories during planar center-out task for two different targets (left and center) for stimulation off (dark grey) and on(blue) condition. The inset block shows the inability of SCS01 to reach to the center target without stimulation **(c)** EMG signals for the left target during reach (light blue highlight) and pull phase (pink highlight) without (dark grey) and with stimulation(blue). **(d)** synergy vector(c) for left target corresponding to the increasing timeseries synergy activation. **(e)** EMG signals for the center target during reach (light blue highlight) and pull phase (pink highlight) for the Center target without (dark grey) and with stimulation(blue). **(f)** Synergy vector for the center target with(blue) and without stimulation (dark grey) for reach (light blue highlight) and pull phase (pink highlight). **(g)** Kinematic trajectories for reaching-out task with(blue) and without (dark grey) stimulation for reach (solid line) and pull phase (dashed line) **(h)** Muscle activity with(blue) and without (dark grey) stimulation during reach(blue highlight) and pull phase(pink highlight) for planar reaching-out task. **(i)** Synergy vector corresponding to the reach (blue highlight) and pull phase (pink highlight) of the movement with(blue) and without(dark grey) stimulation.

**Extended Data Table 1.** Fugl-Meyer Assessment longitudinal breakdown. A breakdown table of the scores for each of the 7 FM-UE assessment categories. In bold, is the total score for the motor function subcategory which is the sum of the Motor Upper Extremity, Motor Wrist, Motor Hand, and Motor coordination/speed sections. The rightmost column indicates the maximum possible score for each category.

|  | Pre-Study | Mid Study | End Study | 1 mo. Post Study | Total Possible |
| --- | --- | --- | --- | --- | --- |
| **SCS01** |  |  |  |  |  |
| Passive Joint Motion | 21 | - | 21 | 21 | 24 |
| Joint Pain | 24 | - | 24 | 24 | 24 |
| Motor Upper extremity | 19 | 20 | 27 | 26 | 36 |
| Motor Wrist | 3 | 3 | 5 | 5 | 10 |
| Motor Hand | 9 | 7 | 11 | 11 | 14 |
| Motor coordination/speed | 4 | 4 | 4 | 4 | 6 |
| **Total Motor Function** | **35** | **34** | **47** | **46** | **66** |
| Sensation | 11 | 11 | 11 | 11 | 12 |
| **SCS02** |  |  |  |  |  |
| Passive Joint Motion | 22 | 20 | 20 | 20 | 24 |
| Joint Pain | 24 | 23 | 23 | 24 | 24 |
| Motor Upper extremity | 11 | 14 | 13 | 12 | 36 |
| Motor Wrist | 0 | 0 | 0 | 0 | 10 |
| Motor Hand | 0 | 0 | 1 | 1 | 14 |
| Motor coordination/speed | 4 | 4 | 4 | 4 | 6 |
| **Total Motor Function** | **15** | **18** | **18** | **17** | **66** |
| Sensation | 2 | 1 | 1 | 2 | 12 |

**Extended Data Table 2.** Modified Ashworth Scale longitudinal breakdown. A breakdown table of the individual MAS scores for each joint tested across all days of the trial. In each case, a score of 0 corresponds to no spasticity, and a score of 4 indicates no mobility at all.

| Study Day | Elb Flx | Dig Flx | Shld ER | Shld IR | Shld ABD | FA Sup | Wr Flx | Pron | Elb Ext |
| --- | --- | --- | --- | --- | --- | --- | --- | --- | --- |
| **SCS01** |  |  |  |  |  |  |  |  |  |
| 1 | 1.5 | 1.5 | 1.5 | 1.5 | 1.5 | 2 | - | - | - |
| 2 | 1.5 | 1 | 1.5 | 1.5 | 1.5 | 1.5 | - | - | - |
| 3 | 1.5 | 1.5 | 1.5 | 1.5 | 1.5 | 1.5 | - | - | - |
| 4 | 1 | 1 | 1.5 | 1.5 | 1.5 | 2 | - | - | - |
| 7 | 1.5 | 1.5 | 1.5 | 1.5 | 1.5 | 1.5 | - | - | - |
| 8 | 1 | 1 | 1.5 | 1.5 | 1 | 2 | - | - | - |
| 9 | 1 | 1.5 | 1.5 | 1.5 | 1 | 2 | - | - | - |
| 10 | 1 | 1 | 1 | 1 | 1 | 1.5 | - | - | - |
| 11 | 1 | 1 | 1.5 | 1 | 1 | 2 | - | - | - |
| 14 | 1 | 1 | 1 | 1 | 1.5 | 1.5 | - | - | - |
| 15 | 1.5 | 1.5 | 1.5 | 1.5 | 1 | 2 | - | - | - |
| 16 | 0 | 1 | 1.5 | 1.5 | 1 | 1.5 | - | - | - |
| 17 | 1 | 1.5 | 1.5 | 1 | 1 | 1.5 | - | - | - |
| 18 | 1 | 1.5 | 1 | 1 | 1.5 | 2 | - | - | - |
| 22 | 1 | 1 | 1.5 | 1.5 | 1.5 | 1.5 | - | - | - |
| 23 | 1 | 1.5 | 1.5 | 1.5 | 1.5 | 2 | - | - | - |
| 24 | 1 | 1.5 | 1 | 1 | 1 | 2 | - | - | - |
| **Post study 52** | 1 | 1.5 | 1.5 | 1.5 | 1 | 1.5 | - | - | - |
| **SCS02** |  |  |  |  |  |  |  |  |  |
| 1 | 2 | 1.5 | 2 | 0 | - | - | 3 | 1.5 | - |
| 2 | 2 | 1.5 | 2 | 1.5 | 2 | - | 3 | 1.5 | - |
| 3 | 2 | 1.5 | 2 | 1 | 2 | - | 3 | 1.5 | 1.5 |
| 4 | 2 | 1.5 | 2 | 1.5 | 1.5 | - | 3 | 2 | 1 |
| 7 | 2 | 1.5 | 2 | 1 | 1.5 | - | 3 | 1.5 | 1.5 |
| 8 | 2 | 2 | 2 | 1.5 | 1.5 | - | 3 | 0 | 0 |
| 9 | 2 | 2 | 1.5 | 0 | 1.5 | - | 3 | 0 | 0 |
| 10 | 2 | 1.5 | 2 | 1.5 | 2 | - | 3 | 2 | 1 |
| 11 | 2 | 2 | 2 | 0 | 1 | - | 3 | 1 | 1 |
| 14 | 2 | 1 | 1.5 | 0 | 1.5 | - | 3 | 0 | 0 |
| 15 | 2 | 1.5 | 2 | 1.5 | 1.5 | - | 3 | 1 | 1 |
| 16 | 2 | 1.5 | 1.5 | 1 | 1.5 | - | 3 | 1.5 | 1 |
| 18 | 2 | 1.5 | 2 | 1.5 | 2 | - | 3 | 2 | 1 |
| 21 | 1.5 | 1 | 1 | 0 | 0 | - | 3 | 1 | 0 |
| 22 | 2 | 1 | 1.5 | 0 | 0 | - | 3 | 1 | 2 |
| 23 | 2 | 1.5 | 1.5 | 1 | 1.5 | - | 3 | 1.5 | 0 |
| 24 | 2 | 1.5 | 2 | 1 | 1 | - | 3 | 1.5 | 1 |

## Methods

### Trial and Subject information

All experimental protocols were approved by the University of Pittsburgh Institutional Review Board (Protocol STUDY19090210) under an abbreviated IDE. The study is published on ClinicalTrial.gov number NCT04512690.

#### Inclusion criteria

Subjects between 21-70 years of age who had suffered from an ischemic or hemorrhagic stroke more than 6 months prior to the start of the study were eligible for participation. All subjects had hemiparesis affecting their upper limb and had a pre-study FM-UE score between 7 and 45. Prior to the study, participants were screened via a medical evaluation. Candidates with severe co-morbidities, previously implanted medical devices, claustrophobia, or who were pregnant, or breastfeeding were excluded from the study. Subjects were not receiving any anti-spasticity, anti-epileptic, or anti-coagulation medications for the duration of the study period.

#### Study protocol

After screening and pre-study baselines, subjects were implanted with clinical spinal cord leads. Starting from day 4 post-implant, subjects underwent experimental assessments 5 times per week, 4 hours per day, for a total of 14 sessions. During sessions, scientific measurements of joint torque, movement kinematics, muscle activity, and performance in robotic tasks and simulated activities of daily living were performed with and without stimulation. The implants were removed by post-implant day 29. Study follow up was performed at least 4 weeks after explant. Each assessment and their associated procedures are described in detail below. Assessments during the first 5 to 7 sessions were focused on designing an optimal stimulation strategy that was then maintained for the remaining sessions.

#### Subject information

In this work, we report results from the first 2 subjects participating in our trial. SCS01 (30-35 years) had a right thalamic hemorrhagic stroke >5 years prior to participation in the study. At the time of her participation in our trial, her post stroke residual was a left-sided spastic hemiparesis for which she was receiving botulinum injections in her biceps, brachioradialis, and pronator teres. Botulinum treatments were suspended starting 6 months prior to the study period and continuing through the end of the study. For SCS01, we included in this work, analysis of 138 isometric force test repetitions at multiple joints (54 stim off and 84 stim on) and 36 planar reaches (18 with SCS and 18 without SCS). We also report the results of simulated activities of daily living and other motor tasks that were performed at least 1 session per week (see **Figure 4)**.

SCS02 (45-50 years) had a right ischemic middle cerebral artery stroke >1 year prior to participation in the study. Her post stroke residual at the time of participation was a left-sided spastic hemiparesis complicated by a left wrist flexion contracture despite treatment with splinting. For SCS02, we included in this work, analysis of 42 isometric force tests repetitions at multiple joints (21 stim off and 21 stim on) and 57 planar reaches (38 with SCS and 19 without SCS) that were obtained across multiple days during the study. We also report the results of simulated activities of daily living and other motor tasks that were performed at least 1 session per week (see **Figure 4)**.

Both subjects successfully completed the protocol with no serious adverse events. SCS01 experienced phlebitis several days after the explant procedure at the end of the study that was resolved with oral antibiotics.

### Surgical Procedure

#### Lead Implant and Explant

General anesthesia was induced using propofol and maintained using sevoflurane and propofol at levels that allowed for reliable somatosensory evoked potential monitoring. Short-acting paralytic was used for intubation, but no additional paralytic was given to facilitate intraoperative monitoring of SCS evoked EMG. Both subjects were placed prone and affixed in a 3-pin Mayfield head holder. The back and neck were prepared and draped in typical sterile fashion and prophylactic antibiotics were administered. A small incision was made over the T1-T2 laminas using fluoroscopic guidance, and the tissue was dissected to expose the fascia. A Tuohy needle was inserted into the T1-T2 epidural interspace and used to guide the placement of a clinically approved 8 contact percutaneous spinal lead (PN 977A260, Medtronic). The first (rostral) lead was threaded rostrally and steered *in situ* using fluoroscopy towards the lateral aspect of the spinal cord such that the most distal contact was positioned at the base of the C3 vertebral body.

To confirm placement of the distal lead and ensure that we could recruit motor pools of the upper arm as proximal as the trapezius, we delivered current controlled monopolar stimulation using an intraoperative neuromonitoring system (Xltek Protektor, Natus Medical). Stimulation pulses were delivered at 1-2 Hz on representative electrodes of the array and we measured compound muscle action potentials (CMAPs) using intramuscular needle electrodes (ipsilateral trapezius, anterior deltoid, medial deltoid, posterior deltoid, biceps, triceps, pronator teres, wrist flexors, wrist extensors, abductor pollicis, and abductor digiti minimi; and contralateral bicep and wrist extensors). We also recorded contralateral activity to ensure that SCS did not induce cross-over effects to the other arm. Once satisfied with the lead placement, the Tuohy needle was removed, and the lead was sutured to the fascia to prevent lead migration.

The second (caudal) lead was placed through the same incision and T1-T2 interspace, this time, ensuring that the most proximal contact was positioned at the T1 vertebral body. As before, intraoperative electrophysiology was performed to ensure proper placement, verifying that SCS could recruit motor pools of the most distal muscles in the hand including abductor pollicis and abductor digiti minimi. Once in final position, the two leads (rostral and caudal) overlapped to provide complete coverage of spinal segments C4 to T1. The distal ends of both leads were tunneled subcutaneously and exited through a separate stab incision over the left flank. Both incisions were closed, and the externalized portion of the leads were covered.

To explant the arrays at the conclusion of the study period, the patients were prepared in a similar fashion to the implantation surgery. The upper thoracic incision was re-opened, and the lead wires were cut and removed proximally. The distal end of the leads were removed through the lateral exit wound and both incisions were closed.

#### Recruitment Curves

To evaluate the specificity of SCS in recruiting individual motor pools, recruitment curves were performed on each of the 16 contacts. Stimulation was delivered at 1-2 Hz on one electrode at a time with gradually increasing current amplitude while simultaneously recording CMAPs from all muscles. The peak-to-peak amplitude of the SCS-induced CMAPs were measured, one for each stimulus amplitude, and normalized to the maximum amplitude recorded on that muscle across all measured trials.

#### Frequency Dependent Suppression

To validate that stimulation was activating dorsal sensory afferent fibers and not directly recruiting ventral motor efferent fibers, we evaluated the stimulation frequency dependent response of CMAP amplitudes across several representative electrodes. Current amplitude was fixed at a level above the motor threshold (the amplitude above which CMAPs were reliably induced). Pulse frequency was then increased from 1-2 Hz up to 20 Hz and the relative, normalized, peak-to-peak amplitude of CMAP responses were compared.

### Medical imaging

#### X-ray Imaging

X-ray images were acquired at weekly timepoints in both axial and sagittal views to ensure the stability of lead position.

#### Lesion Segmentation

MRI was acquired using a 3-T Prisma System (Siemens) using a 64-channel head and neck coil. A T1-weighted structural image was captured using a magnetization-prepared rapid gradient echo (MPRAGE) sequence (TR = 2300 ms; TE = 2.9 ms; FoV = 256 × 256 mm^2^; 192 slices, slice thickness = 1.0 mm, in-plane resolution = 1.0 × 1.0 mm). Lesion segmentation was performed manually for each slice of the sequence using the MRIcron image viewer (NITRC) and the resulting region of interest (ROI) was smoothed on all planes using a gaussian smoothing kernel with a full-width at half-maximum of 2mm. MRIcro_GL (NITRC) was used to visualize and export the resulting segmented overlays.

#### High definition fiber tracking (HDFT)

The same 3-T MRI scanner was configured to use a diffusion spectrum imaging scheme to capture a total of 257 diffusion samples. The maximum b-value used was 4000 s/mm^2^ and the in-plane resolution and slice thicknesses were 2 mm. The accuracy of b-table orientation was examined by comparing fiber orientations with those of a population-averaged template^52^.

The diffusion data were reconstructed in the MNI space using q-space diffeomorphic reconstruction^53^ to obtain the spin distribution function^54^. A diffusion sampling length ratio of 1.25 was used. The output resolution in diffeomorphic reconstruction was 2 mm isotropic. The restricted diffusion was quantified using restricted diffusion imaging^55^. The tensor metrics were calculated and a deterministic fiber tracking algorithm^56^ was used to reconstruct the cortico-spinal tract fibers. A tractography atlas^52^ was used to map left and right cortico-spinal tracts with a distance tolerance of 16 mm. For the fiber tracking, we used: an anisotropy threshold of 0.035, an angular threshold of 50 degrees, and a step size of 1 mm. Tracks with lengths shorter than 10 mm or longer than 200 mm were discarded. A total of 1,000,000 seeds were placed. Topology-informed pruning^57^ was applied to the tractography with 16 iterations to remove false connections. We then calculated the mean fractional anisotropy (FA) values for left and right cortico-spinal tract and the percentage of asymmetry was computed using Stinear’s formula:

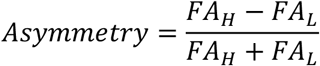

Where *FAL* is the mean FA value of the CST in the lesioned hemisphere and *FAH* is the mean FA value of CST in the intact hemisphere.

### Custom Stimulation Controller

During the trial, SCS was delivered using a clinical grade, single channel, current controlled stimulator (DS8R, Digitimer) and a high-current compliant 1-to-8 multiplexer (D188, Digitimer). Current could be delivered to any contact by connecting it to the multiplexer and selecting the associated output channel. A custom-built microcontroller-based (Arduino Due, Arduino) control unit set pulse timing, amplitude, and output channel for each stimulus. Pulse width, inter-pulse interval, and waveform shape were fixed by the stimulator which ensured proper charge balancing and safe operation. Each pulse was a cathodic-first, biphasic square waveform with 200 us (SCS01) to 400 us (SCS02) monophase pulse width and 10 us inter-pulse interval. Cathodic and anodic phases were equivalent in amplitude and duration.

The control unit triggered each stimulus with a digital trigger pulse and set pulse amplitude using a continuous analog signal between 0-3.3 V. The DS8R hardware was configured for safety such that it could not produce amplitudes higher than 10.23 mA. Despite the stimulator comprising a single current source, the control unit’s firmware enabled semi-synchronous stimulation across multiple channels by rapidly switching the output channel after each pulse (**Extended Data Figure 2 e and 5 and)**. The time between pulses on separate channels was measured to be 2.2 ms, giving enough time for the multiplexer to fully switch output channels. During the study, we used this system to deliver stimulation on up to 4 separate spinal electrodes at up to 100 Hz. All programmable stimulation settings were configurable using a graphical user interface (GUI) developed in MATLAB which communicated with the control unit via a virtual serial port over a USB connection. Stimulation frequency, channel, duration, latency, and amplitude could all be configured manually via the GUI. Each channel could also be set to deliver a single pulse, a pulse train of fixed duration or pulse count, or continuous stimulation **(Extended Data Figure 2)**.

A custom command protocol was implemented to facilitate communication between the GUI and control unit (**Extended Data Figure 2 b and d)**. Communication was always initiated by the GUI with a command packet comprising the length in bytes of the packet, a 1-byte command, and 0-6 bytes of data. Possible commands included triggering or terminating stimulation, clearing the current configuration, reading or writing a parameter, configuring the microcontroller to accept new parameters (program mode), saving parameters, and an initialization handshake. When writing parameters, the length and command bytes were followed by the parameter to be set, the channel (if applicable), and the value to be written. When reading parameters, the data payload comprised only the parameter to be read. All commands were followed by a response packet from the microcontroller comprising the length of the packet, an echo of the command received, a data payload if applicable (for example when reading parameters), and a status byte indicating whether the command was executed correctly.

### EMG Acquisition

To assess muscle activity during movement, surface electromyography (sEMG) was recorded using a wireless EMG system (Trigno, Delsys Inc.). Up to 14 synchronized wireless sensors (Avanti Trigno, Delsys Inc.) were used to amplify, digitize, and wirelessly transmit EMG signals to a base station unit. Each sensor sampled the analog signal at 1925.925 Hz and applied a hardware bandpass filter of 20-800 Hz. Once the signals were received by the base station, they were converted back to an analog waveform and resampled at 2500 Hz by a data acquisition system (PCI-6255, National Instruments) for synchronization with other task events. The Trigno system has a known, fixed wireless latency of 59.6 ms.

At the beginning of each experimental session, the arm and hand were cleaned using isopropyl alcohol. Skin safe adhesive was used to secure the EMG sensors to the subject’s arm. Depending on the muscles of interest for a particular experiment, we recorded from up to 14 individual muscles of the arm and hand; including the trapezius, anterior deltoid, medial deltoid, posterior deltoid, biceps, triceps, pronator teres, wrist flexors, wrist extensors, extensor digitorum, and abductor pollicis, whose locations were identified by palpation while the subject was instructed to perform simple movements. Sensors were then carefully removed at the end of each session.

### Single joint isometric torque

Maximum isometric strength was measured for the shoulder, elbow, and wrist joints (when possible) using a robotic torque dynamometer (HUMAC NORM, CSMi). To measure torque, the robot’s manipulandum was positioned and held at a fixed angle and the subject was asked to apply their maximum force while flexing or extending the joint under test for a sustained period of 5 seconds followed by a 10 to 15 second break. This procedure was repeated 5 times to complete a set. For each joint, the system was configured such that the joint was at a nominal and comfortable angle and so that it was aligned with the manipulandum’s center of rotation. The HUMAC NORM’s suggested configurations were used, when possible, but SCS02 was unable to support the weight of her arm and so was placed in a seated position to measure elbow and shoulder torques instead of the suggested supine position. In addition, a splint was used to secure SCS02’s hand to the manipulandum to assist her in holding the handle securely and a counterweight was used where appropriate to offset the mass of the manipulandum and allow for more sensitive measurements. The maximum torque value within each repetition was considered for analysis.

Grip force was measured using a hand dynamometer. Participants were asked to hold the dynamometer and apply their maximum grasping force for five seconds. Each measurement comprised the highest force produced on each of 3 attempts and data were combined across days to assemble enough data for statistical comparison.

### Planar reach and pull kinematics

To evaluate upper limb motor control during directed reach and pull movements, we used a robotic augmented reality exoskeleton system (KINARM, BKIN Technologies). Participants were secured in a modified wheelchair and their arms were suspended in the exoskeleton to remove the effects of gravity. The platform displayed virtual targets onto a dichroic augmented reality display in front of the subject that allowed them to visualize their hand position relative to the virtual graphics. The robot’s motorized joints permitted the application of a mechanical load to the subject’s movements.

#### Center Out Task

For this task, the participants were asked to reach from a central starting position to one of 3 targets displayed using the AR display, then return to the starting position. On each trial the starting position was displayed, and the robot moved the subject’s arm into position, locking it in place. Next the target was presented, and the exoskeleton was unlocked. An audio cue was played after a randomized 100 to 700 ms delay indicating that the subject could begin their movement. The participant was given 10 (SCS01) or 15 (SCS02) seconds to complete each trial. A target was considered acquired when the subject’s index finger was within a 0.5 cm radius of the target center for 500 ms. An audio cue indicated the end of the reach phase. If the subject was unable to reach the target, the robot returned the arm to the starting position and the next target was presented. If the trial was successful, the subject’s finger was positioned in the center of the target in preparation for the pull phase and locked in place. After a 500 ms delay, the arm was unlocked followed by a final audio cue after another 100-700 ms delay indicating the start of the pull phase, and the subject was required to return their hand to the starting position. In some trials, a load of 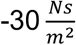 was applied isotropically to the movement using the exoskeleton to increase the task difficulty. Each target was presented 6 times in random order (unless otherwise noted). For each subject, appropriate targets were selected based on their individual range of motion.

The following metrics were calculated for each trajectory to compare kinematic quality. Trajectory smoothness was calculated as the number of peaks in the velocity profile for both the reach and pull phases. We also measured the total time of the combined reach and pull phases. Total path length was calculated and normalized to the Euclidean distance between the starting position and the target; more efficient movements had a lower value. Finally, the variance of each trajectory was calculated as the mean deviation of the actual trajectory from the mean trajectory calculated across all 5 repetitions of the movement.

#### Open-Ended Reaching Task

The subject was presented with 3 equally spaced horizontal lines (approximately 15, 25, and 30 cm away from the participant) and was asked to reach from a starting position to the furthest line they could. In this way we assessed how far the subject could reach in an open-ended manner. During each task, the participant started with their hand as close to their body as they could (maximum elbow flexion). After a verbal cue, they began their movement with the goal of passing the farthest line possible. Once the subject indicated that they had reached their maximum distance, another verbal cue indicated that they should return to their initial position. Task events were manually labeled during the trial by the experimenter. Each set comprised 5 repetitions.

As in the center-out task, a set of metrics was calculated for each trajectory; reach and pull phases were considered separately. Movement duration was calculated as the time it took from the beginning of each phase for the subject to cross the second horizontal line (25 cm) during reach and the first horizontal line during pull (15 cm). Maximum distance was measured as the axial distance between the point closest to the subject and the point furthest from the subject in each phase. Range of motion of the elbow during the task was considered as the angle difference between the most acute and most obtuse elbow angles achieved during each phase. As a metric of smoothness, the number of peaks in the elbow angle velocity profile was counted. Total path length measured the total length of the trajectory from the starting point to the second line (25 cm; reach phase) or from the end position to the first line (15 cm; pull phase) and was normalized by the phase duration. Finally, as a measure of variance, we calculated the distribution of each trajectory timepoint from the mean trajectory. A distribution skewed towards the left indicated that more samples were close to the mean trajectory, whereas a distribution with values towards the right indicated large deviations from the mean trajectory and therefore more variance.

### 3D reaching

#### Fast reaching task

The participant was presented with 6 targets, all axially equidistant from the subject, but at varying heights and lateral positions. The 3 “lower” targets were at table surface height and the 3 “upper” targets were raised to require shoulder flexion beyond 90 degrees. There was a left, center, and right target at each height. A 7^th^ position was placed directly in front of the subject and was used as a “home” position. Starting with their arm outside the working area, the subject was asked to first touch the home position then touch each of the 6 targets, returning to the home position after each target. For this task, we asked the subject to perform the sequence as fast as possible. The total time it took to reach all 6 targets was recorded. For ease of comparison, the average time to acquire one target was calculated from this total.

#### Robotic 3D reaching task

As an alternative to the fast-reaching task, we used an exoskeleton robot (ARMEO POWER, Hocoma) to assist 3D movements when the subject was unable to lift their arm against the force of gravity (SCS02). This robotic system provides motorized support at each joint of the arm and measures kinematic variables in real time allowing for a subject’s real-world movements to be displayed in a virtual video game environment. For this task, objects were presented within a virtual room and the subject was asked to reach toward each object and move it to a different position within the room (ARMEO POWER cleanup game). The robot was configured to provide 50% weight support and assist movements at the “Low Support” setting. Game difficulty was set to “Easy”. Each game lasted 3 minutes and the goal was to move as many objects as possible within the time limit. We then calculated the average time per object to get a comparable measure to the fast-reaching task.

### Clinical Assessments

#### Fugl-Meyer

The Fugl-Meyer Upper-Extremity assessment is a standardized evaluation of upper limb motor control and sensory function^58^. It includes 7 categories of assessments including passive and active range of motion, joint pain, proprioception, and tactile sensation. In total, there are 126 possible points. However, all scores reported in this manuscript correspond to the “Motor Function” sub-score which has a maximum value of 66. A trained medical professional conducted and scored the exam at 4 different timepoints: pre-study, mid-study (approximately 2 weeks after implant), end-of-study (4 weeks), and post-study (1 month after explant).

#### Modified Ashworth Scale

To ensure that SCS was not exacerbating joint spasticity, we performed the Modified Ashworth Scale (MAS) each session day at the beginning of the session. This assessment involves passive manipulation of each joint, and ranking spasticity levels from 0-4 (0 being no spasticity). A trained medical professional performed and scored the assessment each day. Here we report both a full breakdown of all joint scores measured on each day for both subjects as well as a “summary score”. The summary score was taken to be the average score across all joints for each day.

#### Box and Blocks

When possible, we also evaluated the subject’s performance in the “Box and Blocks” task. This is a standardized assessment in which a participant must grasp one small block at a time from one side of a box, lift it over a divider, and drop the block in the other half of the box. The total number of blocks moved from one side to the other within 1 minute was the subject’s score.

### Activities of Daily Living

#### Drawing a spiral

We asked the subject to draw a spiral shape using a marker on a plain piece of white printer paper taped down to a table. The goal of the task was to make the curves as smooth as possible and attempt not to overlap each of the concentric rings. The subject was allowed to comfortably position the pen in their hand using their unaffected hand before starting to draw.

#### Object manipulation

We placed a full, sealed can of soup on a table in front of the participant. The subject was asked to grasp the object from the side, requiring them to supinate their forearm, lift the can, and place it at an adjacent target. This task evaluated the subject’s ability to reach, grasp, lift, and release a moderately heavy object. Here, the subject was not allowed to use their unaffected arm to assist in grasping the object.

In an alternative object manipulation task, we asked the subject to hold a wooden plank with vertical dowels (similar to a tower of Hanoi toy) on their lap using their unaffected hand. We then placed a metal cylinder over one of the dowels. The subject was required to grasp the cylinder, lift it off of the first dowel, align it and place it onto a second dowel, and release the cylinder. An experimenter helped to position the hand on the cylinder before the start of the trial. All other movements were performed by the subject entirely on their own.

#### Lock opening

As a measure of hand dexterity, we positioned a wooden panel with a shackle-style key-actuated lock on a table in front of the subject, who was asked simply to open the lock using their affected limb. To do this task, the participant was required to grasp and stabilize the lock with one hand (e.g. the unaffected hand), use a pinch grip to grasp the key with the other hand (e.g. the affected hand), and supinate the forearm to twist the key and unlock the lock. The subject then removed the lock from its latch on the wooden panel, replaced it by realigning the shank with the latch, and relocked the lock by aligning and pressing the shank back into the body.

#### Self-feeding

The subject was presented with small bite sized portions of food on a plate and a plastic fork. They were tasked with first picking up the fork from a table, using it to secure a piece of food, and perform the complex movement of orienting the food toward their mouth in preparation to eat it. Here, the subject was required to initiate picking up the fork with their affected hand but was allowed to reposition it using their unaffected hand before attempting to pick up the food.

### EMG Analysis

#### Isometric contraction (root mean square analysis)

During isometric contractions, EMG was acquired from appropriate muscles using wireless sensors as described above. Empirically, we observed that deltoid EMG signals contained stimulation artifact during trials where stimulation was active due to the proximity of deltoid muscles to the stimulating electrodes. We removed these artifacts by blanking the signal coinciding with stimulation pulses. All signals were bandpass filtered (25-300 Hz, 5^th^ order Butterworth digital filter) and the root mean square (rms) value was calculated from the filtered data over the full duration of each trial for statistical analysis.

#### Planar reaching (muscle synergy analysis)

Coordinated movements such as reaching and pulling require the timed co-activation of appropriate muscles to produce accurate and controlled limb motion. We measured which muscles were simultaneously active during planar reaching movements by calculating muscle synergies using non-negative matrix factorization (NNMF), a dimensionality reduction technique^59^.

EMG pre-processing was different for SCS01 and SCS02 due to large amplitude stimulation artifacts present in SCS02’s EMG data that were not present for SCS01. For SCS01, stimulation artifact was removed by blanking and the resulting data were bandpass filtered (20-500 Hz, 5^th^ order Butterworth digital filter). For SCS02, EMG were first bandpass filtered using a narrower pass band (10-200 Hz, 5^th^ order Butterworth, digital filter) to remove high frequency components of the stimulation artifact. Notch filters (5^th^ order Butterworth) at 50, 100, and 150 Hz were then used to remove low frequency harmonics of the stimulation artifact. The resulting signals from both subjects were rectified, low-pass filtered (5 Hz, 5^th^ order Butterworth digital filter), and normalized to the maximum EMG value recorded from that muscle over the whole day. Processed EMG was extracted from the reach and pull phases of each movement. Muscle synergies were identified using NNMF.

NNMF decomposes the EMG signals into a synergy activation matrix using the temporal correlation between the activity of individual muscles^59^. The result is a set of one-dimensional timeseries signals for each muscle synergy identified. Each synergy in-turn comprises contributions from multiple muscles as described by a synergy vector. We implemented NNMF with two factors which were selected by observing the point-of-inflection in the residuals vs. number of synergies curve^60^. For each phase of the movement (reach and pull), the primary synergy for that movement was identified as the one that most positively correlated (increased) with the movement. All repetitions of the movement were used to perform the dimensionality reduction. Finally, the contributions of deltoid and elbow muscles were quantified and compared using the primary synergy’s synergy vector.

### Statistics

#### Bootstrapping

All statistical comparisons of means presented in this manuscript were performed using the bootstrap method, a non-parametric approach which makes no distributional assumptions on the observed data. Instead, bootstrapping uses resampling to construct empirical confidence intervals for quantities of interest. For each comparison (e.g. comparing stimulation on vs stimulation off for shoulder torque in SCS01, shown in Figure 1f), we construct bootstraped samples by drawing a sample with replacement from observed measurements, while preserving the number of measurements in each condition. We construct 10,000 bootstrap samples and, for each, calculate the difference in means of the resampled data. A 95% confidence interval for the difference in means is obtained by identifying the 2.5^th^ and 97.5^th^ quantiles for the resulting values. The null hypothesis of no difference in the mean was rejected if 0 was not included in the 95% confidence interval. If more than one comparison was being performed at once, we used a Bonferroni correction by dividing this alpha value by the number of pairwise comparisons being performed.

#### Comparison of distributions

Statistical comparison of distributions was done using a two-sample Kolmogorov-Smirnov (KS) non-parametric test using MATLAB. Again, an alpha value of 0.05 was used. Here, we used this test to compare the variability of kinematic trajectories during 2D planar reaching (the open-ended reaching task). The deviations of each trajectory from the mean trajectory were used to build a distribution of deviations. The resulting distributions for two conditions (stimulation off and stimulation on) could then be compared using the KS test.

